# M-DATA: A Statistical Approach to Jointly Analyzing *De Novo* Mutations for Multiple Traits

**DOI:** 10.1101/2021.05.22.21257421

**Authors:** Yuhan Xie, Mo Li, Weilai Dong, Wei Jiang, Hongyu Zhao

## Abstract

Recent studies have demonstrated that multiple early-onset diseases have shared risk genes, based on findings from *de novo* mutations (DNMs). Therefore, we may leverage information from one trait to improve statistical power to identify genes for another trait. However, there are few methods that can jointly analyze DNMs from multiple traits. In this study, we develop a framework called M-DATA (**M**ulti-trait framework for ***De novo*** mutation **A**ssociation **T**est with **A**nnotations) to increase the statistical power of association analysis by integrating data from multiple correlated traits and their functional annotations. Using the number of DNMs from multiple diseases, we develop a method based on an Expectation-Maximization algorithm to both infer the degree of association between two diseases as well as to estimate the gene association probability for each disease. We apply our method to a case study of jointly analyzing data from congenital heart disease (CHD) and autism. Our method was able to identify 23 genes for CHD from joint analysis, including 12 novel genes, which is substantially more than single-trait analysis, leading to novel insights into CHD disease etiology.

**Author Summary:** Congenital heart disease (CHD) is the most common birth defect. With the development of new generation sequencing technology, germline mutations such as *de novo* mutations (DNMs) with deleterious effects can be identified to aid in discovering the genetic causes for early on-set diseases such as CHD. However, the statistical power is still limited by the small sample size of DNM studies due to the high cost of recruiting and sequencing samples, and the low occurrence of DNMs given its rarity. Compared to DNM analysis for other diseases, it is even more challenging for CHD given its genetic heterogeneity. Recent research has suggested shared disease mechanisms between early-onset neurodevelopmental diseases and CHD based on findings from DNMs. Currently, there are few methods that can jointly analyze DNM data on multiple traits. Therefore, we develop a framework to identify risk genes for multiple traits simultaneously for DNM data. The new method is applied to CHD and autism as a case study to demonstrate its improved power in identifying risk genes compared with single-trait analysis. Our results lead to new insights on the disease etiology of CHD, and the shared etiological mechanisms between CHD and autism.

## Introduction

Congenital heart disease (CHD) is the most common birth defect. It affects 0.8% of live birth and accounts for one-third of all major congenital abnormalities [1, 2]. CHD is associated with both genetic and environmental factors [2]. It is genetically heterogenous and the estimated heritability in a Danish twin study is close to 0.5 [3].

Studies on *de novo* mutations (DNMs) have been successful in identifying risk genes for early on-set diseases as DNMs with deleterious effects have not been through natural selection. By conducting whole-exome sequencing (WES) studies for parent-offspring trios, there are cumulative findings of potential risk genes for CHD and neurodevelopmental disorders by identifying genes with more DNMs than expected by chance [4-6]. However, the statistical power for identifying risk genes is still hampered by the limited sample size of WES due to its relatively high cost in recruiting and sequencing samples, as well as the low occurrence of DNMs given its rarity.

Meta-analysis and joint analysis are two major approaches to improve the statistical power by integrating information from different studies. Meta-analysis studies on WES DNMs and Genome-wide Association Studies (GWAS) for multiple traits have been conducted [7, 8]. However, these approaches may overlook the heterogeneity among traits, thus hinder the ability to interpret finding for each single trait. By identifying the intersection of top genes from multiple traits, some recent studies have shown that there are shared risk genes between CHD and autism [9, 10]. Shared disease mechanism for early-onset neurodevelopmental diseases has also been reported [11, 12]. Based on these findings, joint analysis methods have been proposed and gained success in GWAS and expression quantitative trait loci (eQTL) studies. Studies have shown that multi-trait analysis can improve statistical power [13-19] and accuracy of genetic risk prediction [20-22]. Currently, there lacks joint analysis methods to analyze DNM data on multiple traits globally, with the exception of mTADA [23].

In addition to joint analysis, integrating functional annotations has also been shown to improve statistical power in GWAS [15, 24] and facilitate the analysis of sequencing studies [25] [26]. There is a growing number of publicly available tools to annotate mutations in multiple categories, such as the genomic conservation, epigenetic marks, protein functions and human health. With these resources, there is a need to develop a statistical framework for jointly analyzing traits with shared genetic architectures and integrating functional annotations for DNM data.

In this article, we propose a **M**ulti-trait ***D****e novo* mutation **A**ssociation **T**est with **A**nnotations, named M-DATA, to identify risk genes for multiple traits simultaneously based on pleiotropy and functional annotations. We demonstrate the performance of M-DATA through extensive simulation studies and real data examples. Through simulations, we illustrate that M-DATA is able to accurately estimate the proportion of disease-causing genes between two traits under various genetic architectures. M-DATA outperformed single-trait approaches and methods even if annotation information was not used. Annotations can further boost the power of M-DATA. We applied M-TADA to identify risk genes for CHD and autism. There are 23 genes discovered to be significant for CHD, including 12 novel genes, bringing novel insight to the disease etiology of CHD.

## Methods

### Probabilistic model

First, we consider the simplest case with only one trait, and then we extend our model to multiple traits. We denote *Y*_*i*_ as the DNM count for gene *i* in a case cohort, and assume *Y*_*i*_ come from the mixture of null (*H*_0_), and non-null (*H*_1_), with proportion *π*_0_ =1 − *π* and *π*_1_ = *π* respectively. Let *Z*_*i*_ be the latent binary variable indicating whether this gene is associated with the trait of interest, where *Z*_*i*_ *=0* means gene *i* is unassociated (*H*_0_), and *Z*_*i*_ =1 means gene *i* is associated (*H*_1_). Then, we have the following model:

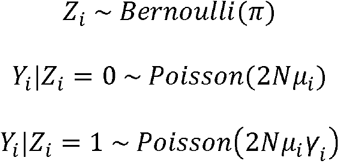

where N is the sample size of the case cohort, *µ*_*i*_ is the mutability of gene *i* estimated using the framework in Samocha, Robinson (27), and *γ*_*i*_ is the relative risk of the DNMs in the risk gene and is assumed to be larger than 1. The derivation of the parameter of the Poisson distribution is the same as that in TADA [6, 28]. We define this model as the single-trait model without annotation in our main text.

To leverage information from functional annotations, we use an exponential link between *γ*_*i*_ and *X*_*i*_,

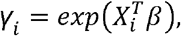

where 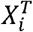 is the transpose of the functional annotation vector of gene *i*, and *β* is the effect size vector of the functional annotations. Under the assumption that risk genes have higher burden than non-risk genes, we expect the estimated value of *γ*_*i*_ to be larger than 1.

Now we extend our model to consider multiple traits simultaneously. To unclutter our notations, we present the model for the two-trait case. Suppose we have gene counts *Y*_*i*1_ and *Y*_*i*2_ for gene *i* from two cohorts with different traits. Similarly, we introduce latent variables *Z*_*i*_ = [*Z*_*i*00_, *Z*_*i*10_, *Z*_*i*01_, *Z*_*i*11_] to indicate whether gene *i* is associated with the traits. Specifically, *Z*_*i*00_ =1 means the gene *i* is associated with neither trait, *Z*_*i*10_ = 1 means that it is only associated with the first trait, *Z*_*i*01_ =1 means that it is only associated with the second trait, and *Z*_*i*11_ =1 means that it is associated with both traits. Then, we have:

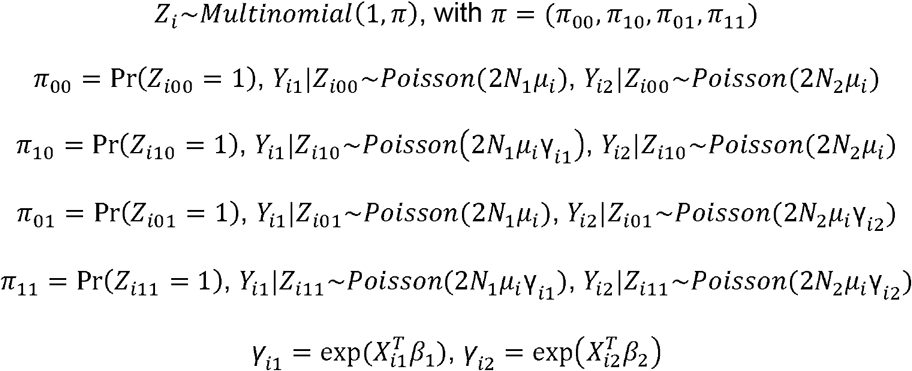

where *π* is the corresponding risk proportion of genes belonging to each class, with ∑_*l* ∈ {00,10,01,11}_ *π*_*l*_ = 1. Then, the risk proportion of the first trait and second trait is *π*_10_ + *π*_11_ and *π*_01_ + *π*_11_, respectively. When there is no pleiotropy of the two traits, *π*_11_ = (*π*_10_ + *π*_11_)(*π*_01_ + *π*_11_). The difference between *π*_11_ and (*π*_10_ + *π*_11_)(*π*_01_ + *π*_11_) reflects the magnitude of global pleiotropy between the two traits. *µ*_*i*_ is the same as our one-trait model. *N*_1_, *γ*_*i*1_ and *X*_*i*1_ are the case cohort size, relative risk and annotation vector of gene *i* for the first trait. *N*_2_, *γ*_*i*2_ and *X*_*i*2_ are similarly defined for the second trait.

Denote *Θ =* (*π, β*_1_, *β*_2_) the parameters to be estimated in our model. As we only consider *de novo* mutations, they can be treated as independent as they occur with very low frequency. The full likelihood function can be written as

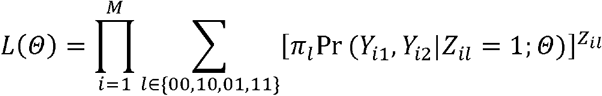

where M is the number of genes. The log-likelihood funciton is

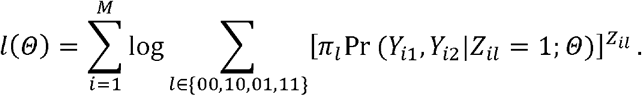

### Estimation

Parameters of our models can be estimated using the Expectation-Maximization (EM) algorithm [32]. It is very computationally efficient for our model without annotation because we have explicit solutions for the estimation of all parameters in the M-step.

By Jenson’s inequality, the lower bound *Q*(Θ) of the log-likelihood function is

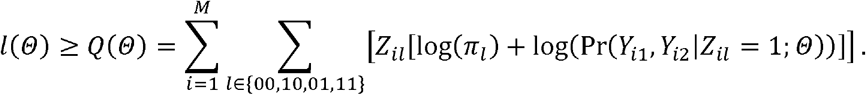

The algorithm has two steps. In the E-step, we update the estimation of latent variables *z*_*il*_, *l* ∈ {00,01,10,11} by its posterior probability under the current parameter estimates in round s. That is,

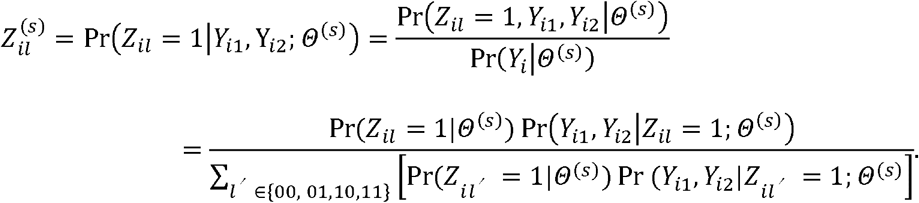

In the M-step, we update the parameters in *Θ* based on the estimation of *z*_*il*_ in the E-step by maximizing *Q*(*Θ*). For *π*, there is an analytical solution, which is

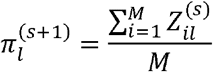

For the rest of derivation, we take the estimation process for the first trait as an example. Taking the first order derivative of *Q*(Θ) with respect to *β*_1_ as 0, we have

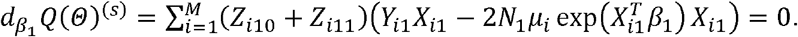

If we do not add any functional annotations to our model (*X*_*i*1_ degenerates to 1 and *β*_1_ degenerates to a scalar), there exists an analytical solution for *β*_1_.

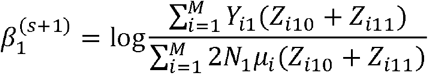

However, there is no explicit solution for *β*_1_, so we adopt the Newton-Raphson method for estimation after adding functional annotations into our model. The second-order derivatives for *Q*(*Θ*) is

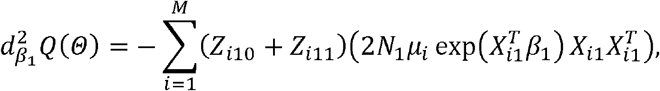

Then, the estimate of *β*_1_ can be obtained as

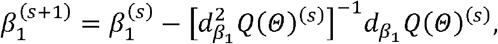

### Functional Annotation and Feature Selection

As we have discussed, there are multiple sources of functional annotations for DNMs. For gene-level annotations, we can directly plug into our gene-based model. For variant-level annotations, it is important to collapse the variant-level information into gene-level without diluting useful information. Simply pulling over variant-level annotations of all base pairs within a gene may not be the best approach. To better understand the relationship, we calculate the likelihood ratio of the DNM counts under *H*_1_ and *H*_0_. Under *H*_1_, for all positions t within a gene *i*, the DNM count *Y*_*it*_ follows the Poisson distribution with relative risk *γ*_*it*_ and mutability *μ*_*it*_, then we have

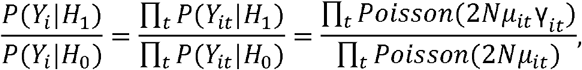

where _*γit*_ = exp(*β*_0_ + *β*_1_*X*_*it*_). There is likely to be at most one mutation at each position *t* due to the low frequency of DNM. We can further simplify the above equation to

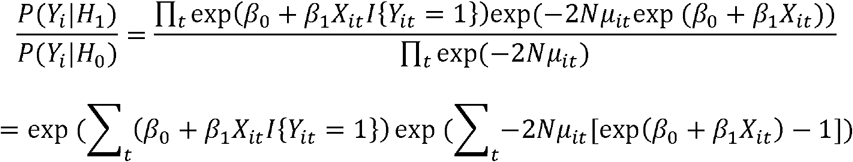

Assuming the variant-level effect size *β*_1_ is small, we can apply Taylor expansion to the second term of the above equation,

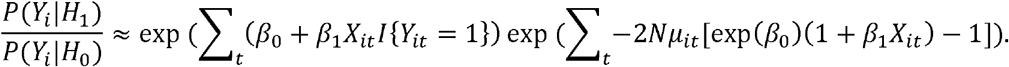

If we center the collapsed variant-level annotations, we can apply ∑_*t*_ *X*_*it*_ =0 to the above equation and further simplify it as

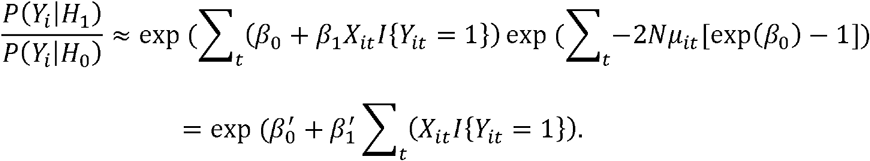

The above approximation motivates us to aggregate variant-level annotations to gene-level annotations by summing up all annotation values of the mutations within a gene after preprocessing each variant-level annotation.

We used variant-level annotations from ANNOVAR [29] in our analysis. We define loss-of-function (LoF) as frameshift insertion/deletion, splice site alteration, stopgain and stoploss predicated by ANNOVAR, and define deleterious missense variants (Dmis) predicted by MetaSVM [30]. Specifically, we included four categories of features including variant-level deleteriousness (PolyPhen (D), PolyPhen(P) [33], MPC [34], CADD [35], REVEL [36], and LoF), variant-level allele frequencies (gnomAD_exome and gnomAD_genome [31]), variant-level splicing scores (dbscSNV_ADA_score, dbscSNV_RF_score [37] and dpsi_zscore [38]) and gene conservation scores (pLI and mis_z) downloaded from gnomAD v2.1.1 [31] in real data analysis. To construct gene-level annotation scores, variant-level annotations were collapsed by summing up values calculated from the mutation information for each gene. All continuous gene-level features were normalized before model fitting.

Before performing multi-trait analysis, features were selected separately for each trait by single-trait analysis. For each trait, all gene-level features were evaluated by Pearson’s correlation. If the Pearson’s correlation between two annotations was larger than 0.7, only one annotation was kept. After model fitting, we kept annotations with the absolute values of effect sizes larger than 0.01 and refit the model with the selected annotations. For multi-trait analyses, we constructed the annotation matrices using the features selected from each trait (see more details in the S1 Text.)

### Hypothesis Testing

Without loss of generality, we take the first trait as an example to illustrate our testing procedure. After we estimate the parameters, genes can be prioritized based on their joint local false discovery rate (Jlfdr) [39]. For joint analysis of two traits, the Jlfdr of whether gene is associated with the first trait is

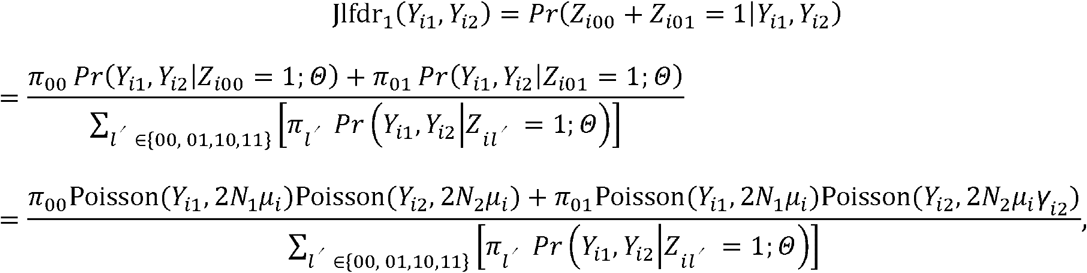

where 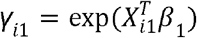 and 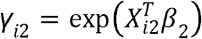. When there is no annotation, both *β*_1_ and *β*_2_ degrade from vectors to single intercept values. Then *γ*_*i*1_ and *γ*_*i*2_ share the same values exp(*β*_1_) and exp(*β*_2_) across all genes. Same formula can be used to compute the Jlfdr of each gene. The definition of the Jlfdr is the posterior probability of a null hypothesis being true, given the observed DNM count vector (*Y*_1_, *Y*_2_). If we consider the first trait, the corresponding null hypothesis is the gene *i* associates with both traits or only associates with the second trait, i.e., Z_i00_ + Z_i01_ = 1. And the corresponding Jlfdr is Jlfdr_1_(Y_i1_, Y_i2_) = Pr (*z*_*i*00_ + *z*_*i*01_ = 1|*Y*_*i*1_, *Y*_*i2*_). In comparison, the *p*-value is defined as the probability of observing more extreme results given the null hypothesis being true, i.e., *p*-value= Pr (More extreme than (*Y*_*i*1_, *Y*_*i*2_)|z_*i*00_ + z_*i*01_ = 1). To compute it, we need to firstly define a partial order for comparing two-dimensional vector (*Y*_1_, *Y*_2_), with which the genes associate with the first trait can stand out. One way to define the partial order is to summarize the vector into a one-dimensional test statistic. Since this is not our focus, we will not discuss how to derive a new test statistic in the article. Although the Jlfdr_1_ already informs the probability of whether the gene is associated with the first trait, we should not directly use it as the p-value to infer the association status due to their different definitions and properties.

The following relationship between Jlfdr and false discovery rates (Fdr) was shown in Jiang and Yu (39),

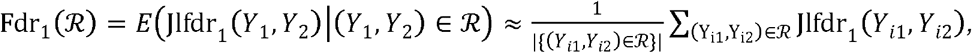

where the rejection region is the set of two-dimensional vector (*Y*_1_, *Y*_2_) such that the null hypothesis can be rejected based on a specific rejection criterion. For example, we can specify a rejection criterion to select genes with large values of the weighted average DNM counts:0.9*Y*_1_ + 0.1*Y*_2_ 2 ≥ 5, then the corresponding rejection region is the upper right region above the line of 0.9 *Y*_1_ + 0.1 *Y*_2_ = 5. Here we omit the gene indicator *i* since the rejection region is defined on DNM count pairs of two traits regardless of the exact gene labels. Jiang and Yu (56) showed that the most powerful rejection region for a given Fdr level q is {Jlfdr_1_(*Y*_1_, *Y*_2_) ≤ *t*(*q*)}.

To determine the threshold *t*(*q*), we sort the calculated Jlfdr_1_value of each gene in an ascending order first. Denote the a-th Jlfdr_1_ value as 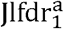. We can approximate the Fdr of the region 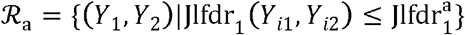 as

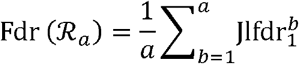

Denote *c* = max {*a* |Fdr (ℜ_a_) ≤q}, and then the threshold *t*(*q*) for Jlfdr_1_ is 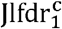. For testing association with the first trait, we reject all genes with Jlfdr_1_(Y_i1_, Y_i2_) ≤*t*(*q*). For both simulation and real data analyses, the global Fdr is controlled at *q* = 0.05. The global Fdr is abbreviated as FDR in the following text.

### Implementation of mTADA

We used extTADA [11] to estimate the hyperpriors input for mTADA. For simulation and real data application, we applied 2 MCMC chains and 10,000 iterations as recommended by the authors [23]. We applied PP>0.8 as the threshold for risk gene inference. We benchmarked the computational time of mTADA and M-DATA on Intel Xon Gold 6240 processors (2.6GHZ).-

#### Misspecified Model

We tested if M-DATA have proper power when functional annotations affect the latent variables z_*il*_, *l* ∈ {00,01,10,11} rather than the relative risk parameters *γ*_*i*1_ and *γ*_*i*2_. Further, we assumed that the latent variable *z*_*i*10_ is associated with the functional annotation vector *X*_*i*1_, which is the functional annotation vector for gene *i* of the first trait, *z*_*i*01_ is associated with *X*_*i*2_, which is the functional annotation vector for gene *i* of the second trait, and *z*_*i*11_ is associated with both *X*_*i*1_ and *X*_*i*2_ through the following forms:

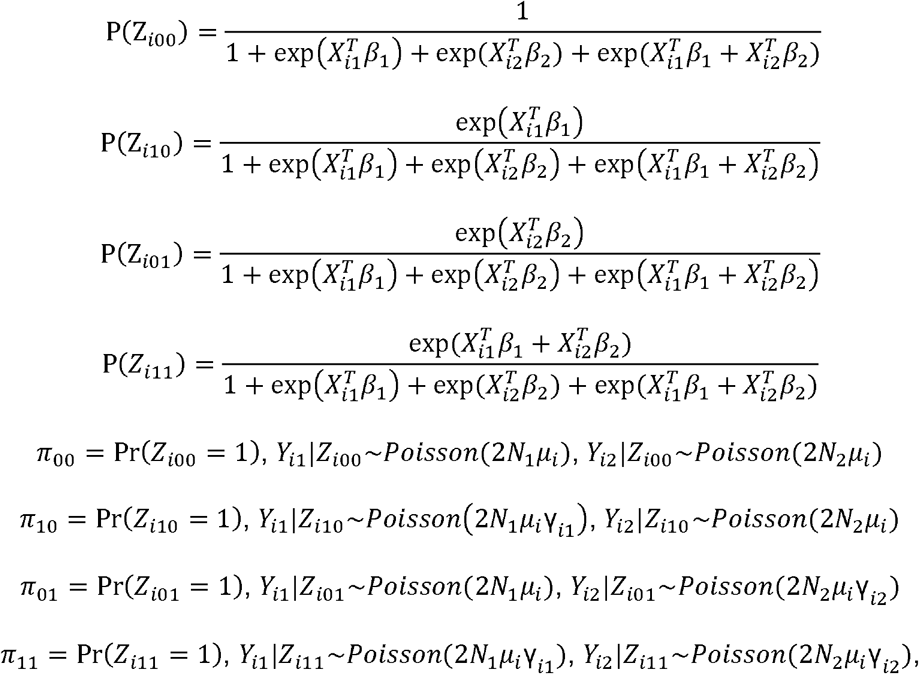

where *π* is the corresponding risk proportion of genes belonging to each class, with ∑_*l* ∈ {00,10,01,11}_ *π*_*l*_ = 1. Here, *μ*_*i*_ is the mutability of gene *i. N*_1_, *γ*_*i*1_ and *X*_*i*1_ are the case cohort size, relative risk and annotation vector of gene for *i* the first trait. Similarly, *N*_2_, *γ*_*i*2_ and *X*_*i*2_ are defined for the second trait.

### Verification and Comparison

#### Estimation Evaluation

We conducted comprehensive simulation studies to evaluate the estimation and power performance of M-DATA. We set the total number of genes M to 10,000, where genes were randomly selected from gnomAD v2.1 [31]. We set the size of the case cohort at 2000, 5000 and 10000, corresponding to a small, medium and large WES study. We assumed the proportion of risk genes to be 0.1 for each trait (i.e., *π*_10_ + *π*_11_ = *π*_01_ + *π*_11_ = 0.1), and varied the shared risk proportion *π*_11_ at 0.01, 0.03, 0.05, 0.07 and 0.09. When *π*_11_ = 0.01, it corresponds to the absence of pleiotropy between two traits, and we expect our multi-trait models to perform similarly as our single-trait models.

We first evaluated the performance of estimation for our models, and then we conducted power analysis for our single-trait models and multi-trait models. To evaluate the estimation performance for multi-trait models, we simulated the true model with two Bernoulli annotations, and set the parameter of the Bernoulli distributions to 0.5 for both traits. We varied the effect sizes of annotations (*β*_*j*0_,*β*_*j*1_,*β*_j2_), *j* = 1,2 from (3,0.1,0.1) (3,0.1,0) and (3,0,0), which corresponds to the cases when both annotations are effective, only the first annotation is effective and no annotation is effective. We evaluated the estimates of shared proportion of risk genes *π*_11_ and the risk gene proportion for a single trait. There are in total 27 simulation settings for estimation evaluation. To obtain an empirical distribution of our estimated parameters, we replicated the process for 50 times for each setting. We simulated the two traits in a symmetrical way, so we only present the results of the first trait. The performance of estimation under the scenario that both annotations are effective((*β*_*j*1_,*β*_*j*2_) = (0.1,0.1), *j* = 1,2) are shown in Fig 1. The rest of scenarios are shown in Fig A in the S1 Text.

**Fig 1.**
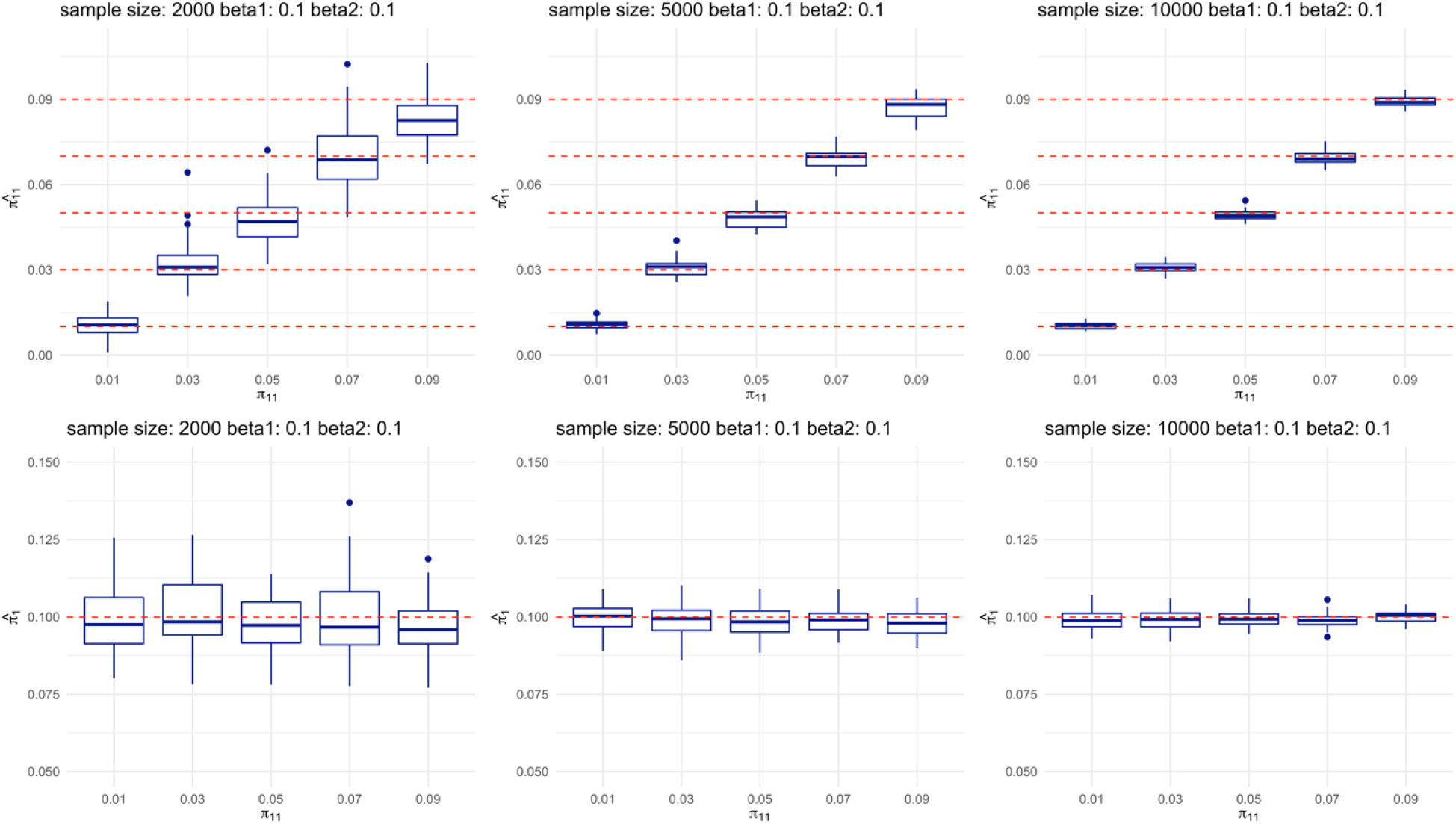
Multi-trait analysis can accurately estimate the proportion of shared risk genes and single-trait risk genes. Top panels show the estimation of shared risk proportion, and bottom panels show the estimation of a single trait. For each panel, each plot from left to right represents study sample size of 2000, 5000, and 10000, respectively. Within each plot, boxes from left to right represent the proportion of shared risk genes being 0.01, 0.03, 0.05, 0.07 and 0.09, respectively. Each scenario is replicated for 50 times in our simulations. True values are shown in red dashed lines.

#### Power Evaluation

Given that the effective number of functional annotations for DNM data in real world is unknown, we explored the power performance of single-trait and multi-trait models when annotations are only partially observed. We varied the effect size of annotations from and, which corresponds to the cases when effect of annotations is weak, moderate, and strong. We assumed that only the first two annotations can be observed. We first demonstrated our model can control FDR (Fig B in the S1 Text) under theses settings and then evaluated power (Fig 2), type I error (Fig C in the S1 Text), and AUC (Fig D in the S1 Text) for our single-trait models and multi-trait models. There are in total 45 simulation settings. Under each setting, the data were simulated based on our multi-trait model with annotations (Methods).

**Fig 2.**
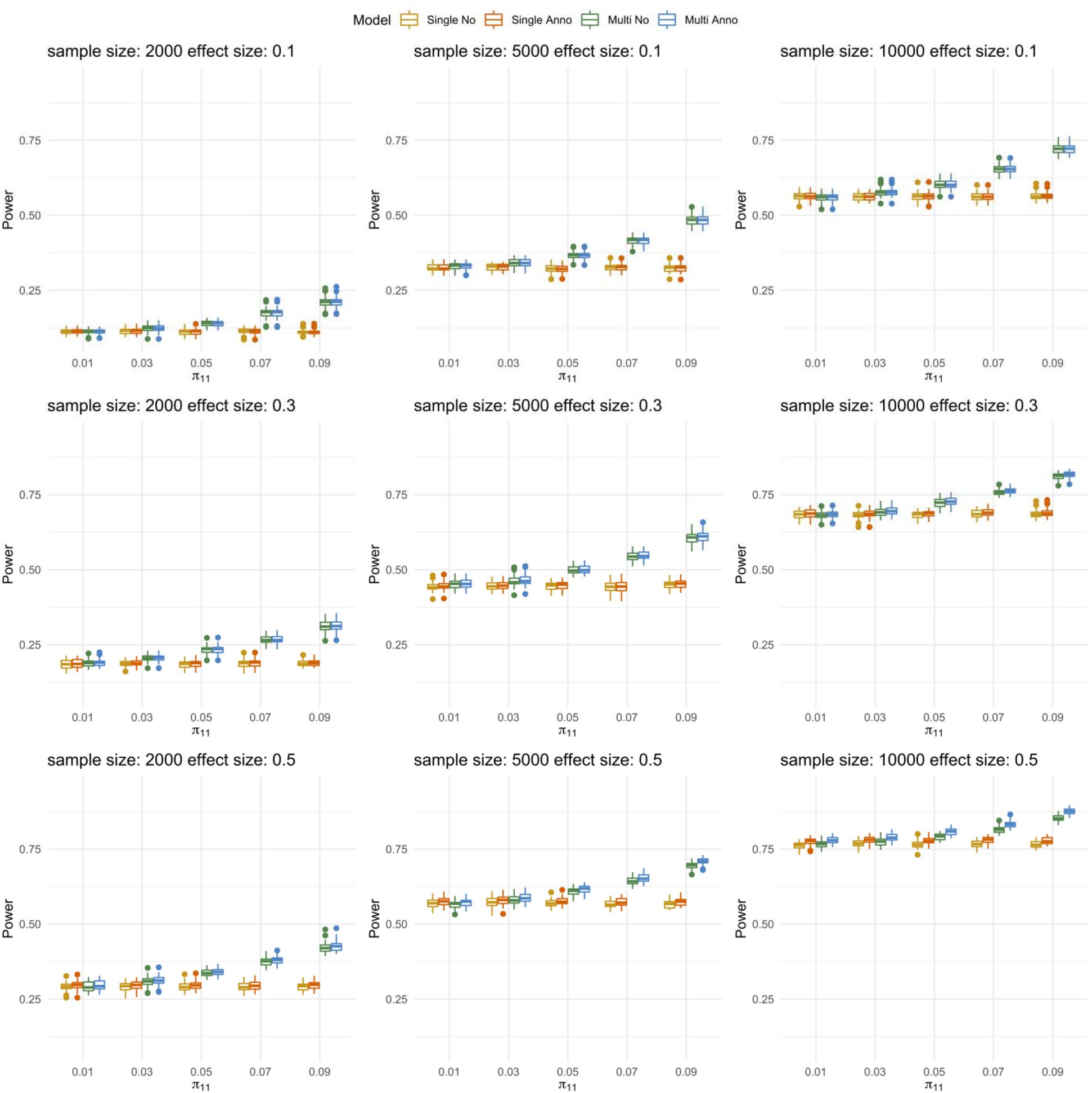
Power performance under different strengths of annotations. The panels from top to bottom show the power performance under weak, moderate and strong annotations, respectively. For each panel, each plot from left to right represents study sample size of 2000, 5000, and 10000, respectively. Within each plot, boxes from left to right represent the proportion of shared risk genes being 0.01, 0.03, 0.05, 0.07 and 0.09, respectively. Each scenario is replicated for 50 times in our simulations.

With the increase of the sample size, the performance of all four models becomes better. Under weak annotations, the power performance of models with annotations and without annotations are comparable. However, when annotations are stronger, the power performance of models with annotations are better than models without annotations (Fig E and Fig F in the S1 Text). With the increase of shared risk proportion, the power performance of multi-trait models become better than single-trait models.

### Comparison with mTADA

Under the same settings in the previous section, we compared the power performance of mTADA and M-DATA. The sample size of the DNM cohort was set as 5000 for both traits. In the simulation, we observed that both methods could control FDR, while mTADA was more conservative than M-DATA for FDR control (Fig G in the S1 Text). M-DATA has higher power than mTADA when the effect size of annotations is larger (Fig 3). The result is consistent with our observation in the real data (Application). In the time comparison, we observed that our method converged faster than the MCMC method adopted by mTADA (Table D in the S1 Text).

**Fig 3.**
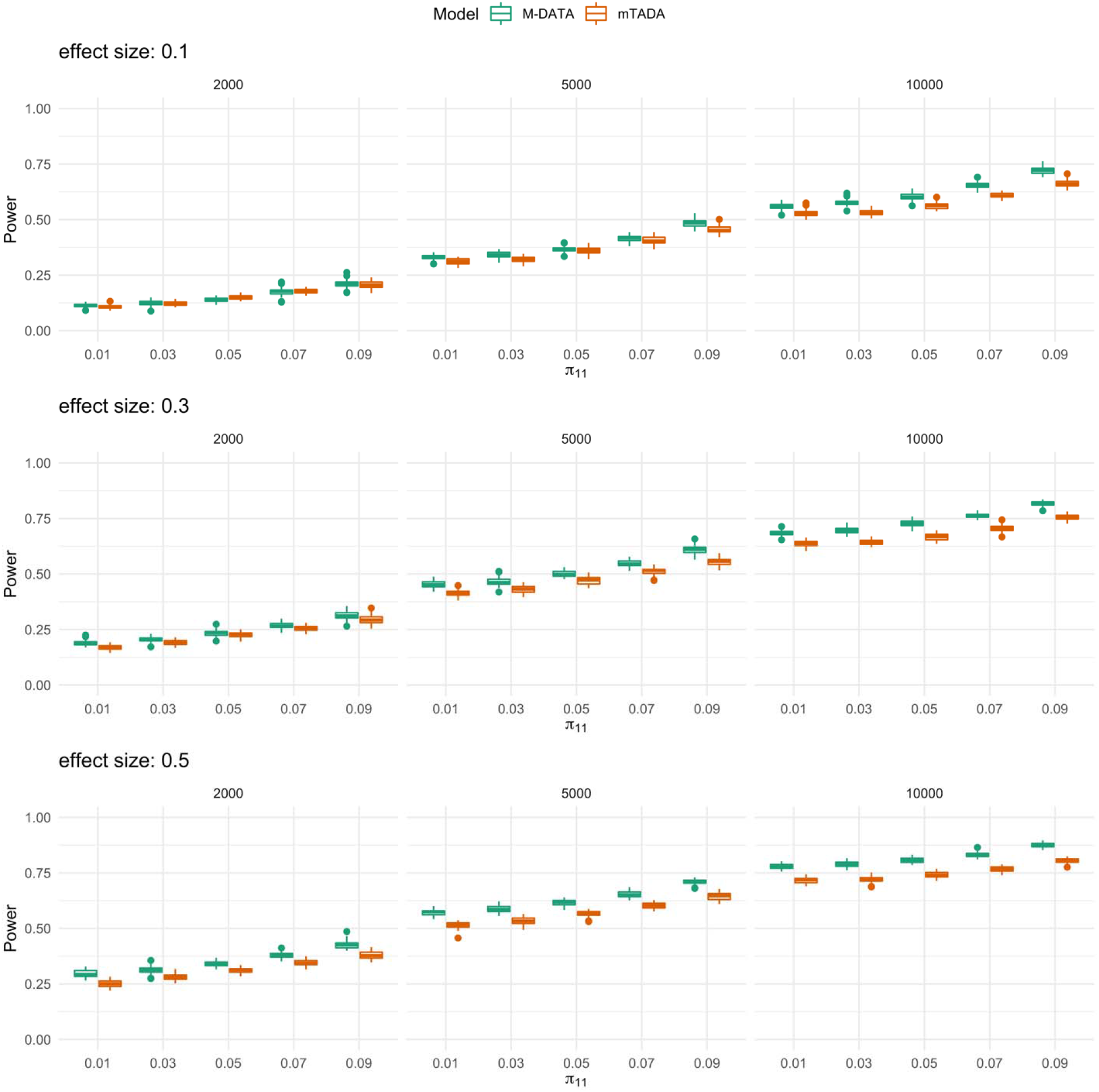
Comparisons of M-DATA and mTADA under different strengths of annotations. The panels from top to bottom show the power performance under weak, moderate and strong annotations, respectively. For each panel, each plot from left to right represents study sample size of 2000, 5000, and 10000, respectively. Within each plot, boxes from left to right represent the proportion of shared risk genes being 0.01, 0.03, 0.05, 0.07 and 0.09, respectively. Each scenario is replicated for 50 times in our simulations.

### Robustness to Model Misspecification

We also evaluated the power performance of M-DATA under misspecified models (Methods), where we simulated two Bernoulli annotations that affect the latent variables *z*_*il*,_ *l* ∈ {00,10,01,11}, and set the parameter of the Bernoulli distributions to 0.5 for both traits. We varied the effect sizes of annotations on the latent variables (*β*_*j*0_,*β*_*j*1_,*β*_*j*2_), *j* 1 = 1,2 at (−3,0.5,0.5), (−3,1,1) and (−3,1.5,1.5), which corresponds to the case when the effect of annotations is weak, moderate, and strong, respectively. The relative risk parameters *γ*_i1_ and *γ*_i2_ were set at 25. We simulated DNM counts under this misspecified model and evaluated the performance of M-DATA multi-trait models for different sizes of DNM cohort (1000, 2000, and 4000). We observed that M-DATA can control FDR under all settings and the multi-trait model with annotations had better power than the multi-trait model without annotation with the increase of the effect size of annotations (Fig 4).

**Fig 4.**
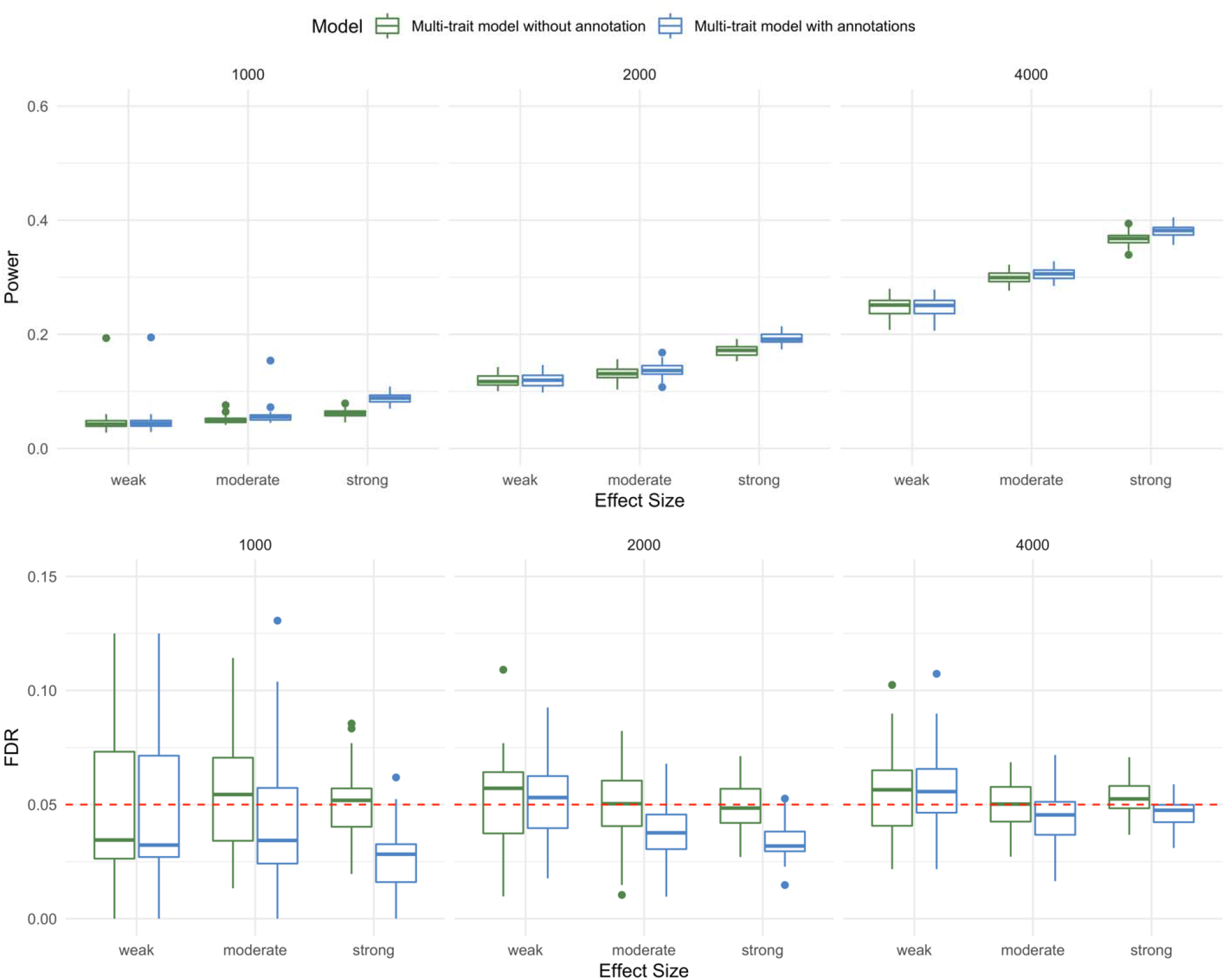
Power and FDR of M-DATA under model misspecification. The top panel and bottom panel show the power and FDR under weak, moderate and strong annotations on the latent variables respectively. For each panel, each plot from left to right represents study sample size of 1000, 2000, and 4000, respectively. Each scenario is replicated for 50 times in our simulations.

## Application

We applied M-DATA to real DNM data from 2,645 CHD probands reported in Jin et al. [4] and 5,623 autism probands acquired from denovo-db [40]. We only considered damaging mutations (LoF and Dmis) in our analysis as the number of non-deleterious mutations is not expected to provide information to differentiate cases from controls biologically [41]. Details of functional annotation and feature selection are included in Methods and S1 Text. In total, there were 18,856 genes tested by M-DATA.

We performed single-trait analysis on CHD and autism data separately, followed by joint analysis both CHD and autism data with the multi-trait models. We compared the performance of single-trait models and multi-trait models for CHD under different significance thresholds. With a stringent significance threshold (i.e., FDR < 0.01), single-trait model without annotation identified 8 significant genes, single-trait model with annotation identified 10 significant genes, multi-trait model without annotation identified 11 significant genes, and multi-trait model with annotation identified 14 genes. With FDR < 0.05, single-trait model without annotation identified 15 significant genes, single-trait model with annotation identified 19 significant genes, multi-trait model without annotation identified 18 significant genes, and multi-trait model with annotation identified 23 significant genes (Table 1). It demonstrates that M-DATA is able to identify more genes by jointly analyzing multiple traits and incorporating information from functional annotations. We visualized the identified genes with Venn diagrams (Fig 5 and Fig H in Text S1).

**Table 1.**
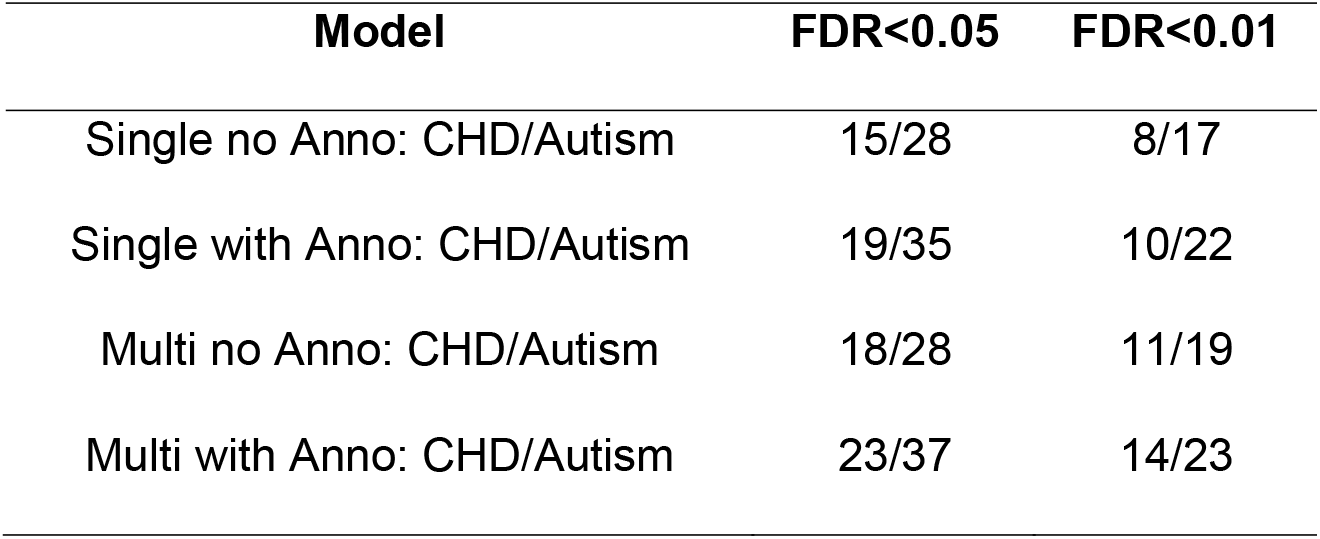
Results for M-DATA Single-Trait and Multi-Trait Models.

| Model | FDR<0.05 | FDR<0.01 |
| --- | --- | --- |
| Single no Anno: CHD/Autism | 15/28 | 8/17 |
| Single with Anno: CHD/Autism | 19/35 | 10/22 |
| Multi no Anno: CHD/Autism | 18/28 | 11/19 |
| Multi with Anno: CHD/Autism | 23/37 | 14/23 |

**Fig 5.**
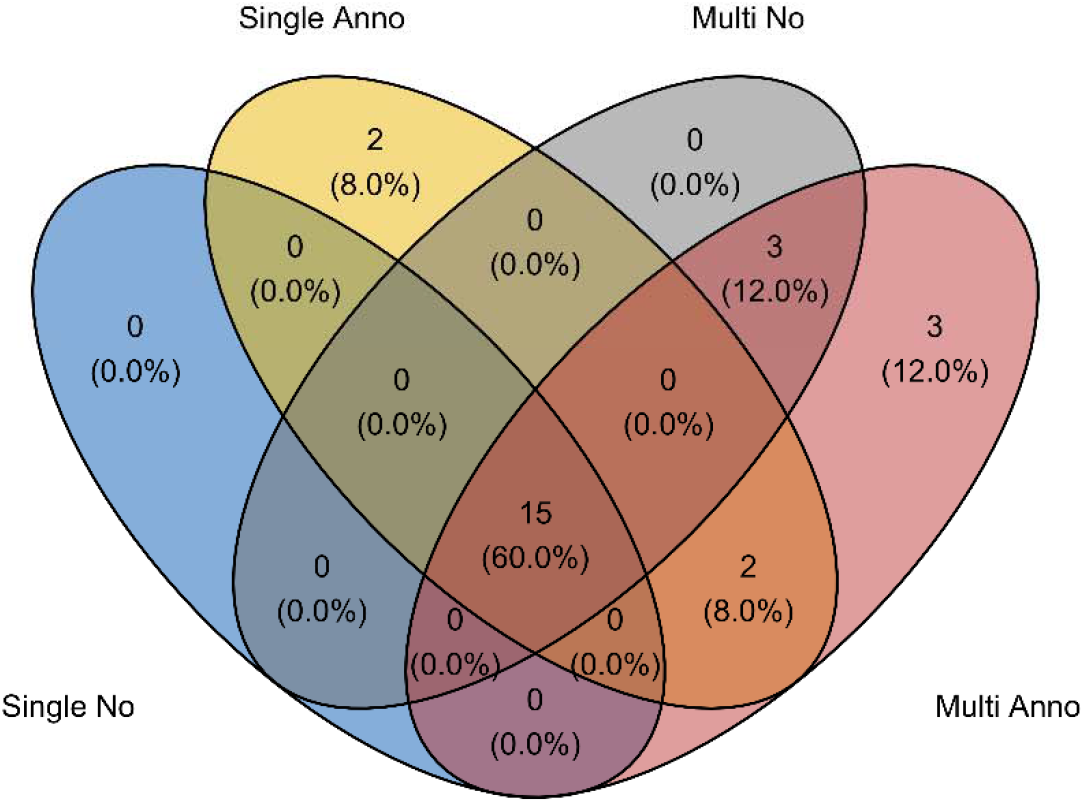
Venn diagram of identified genes in different models. Compared to the single-trait model without annotation, the single-trait model with annotations identified 4 additional genes. Compared to the multi-trait model without annotation, the multi-trait model with annotations identified 5 additional genes. In total, the multi-trait models identified 6 different genes compared to the single-trait models, including 4 novel human CHD genes (*CDK13, FRYL, LZTR1* and *NAA15*).

We further demonstrate the results by taking CHD as an example. Compared with the single-trait model without annotation, the multi-trait model without annotation identified 3 additional genes, which are *FRYL, NAA15* and *PTEN*. Compared with the single-trait model with annotations, the multi-trait model with annotations identified 6 additional genes, including *CDK13, FRYL, LZTR1, NAA15, PTEN* and *RPL5*. There are two additional genes identified by the single-trait model with annotations, but not the multi-trait models. Both of these two genes did not have DNMs in autism and are around the margin of FDR threshold (0.05) for the multi-trait model with annotations (*AHNAK* 0.056, *MYH6* 0.061).

To further illustrate the gain of power from multi-trait analysis, we visualized the posterior probability of being shared risk gene for CHD and autism of identified genes in the multi-trait model with annotations in Fig 6A (CHD) and Fig I in the S1 Text (autism). In the main text, we further illustrate the results with the 23 significant CHD genes. All genes identified by the multi-trait models are annotated with gene names, and the 6 additional genes identified by multi-trait analyses are colored red. From this figure, we can see that most genes (5/6) have high posterior probability of being shared. *RPL5* is at the margin of FDR threshold in the single-trait models and may be prioritized in the multi-trait models by chance (Fig 6B). In addition, we checked the correlation between the FDR of top genes identified by the multi-trait model with annotations in the single-trait model with annotations (Fig 6B). All 6 genes have low FDR (<0.2) in the single-trait model with annotations, which indicates multi-trait analysis can prioritize marginal signals in single-trait analysis.

**Fig 6.**
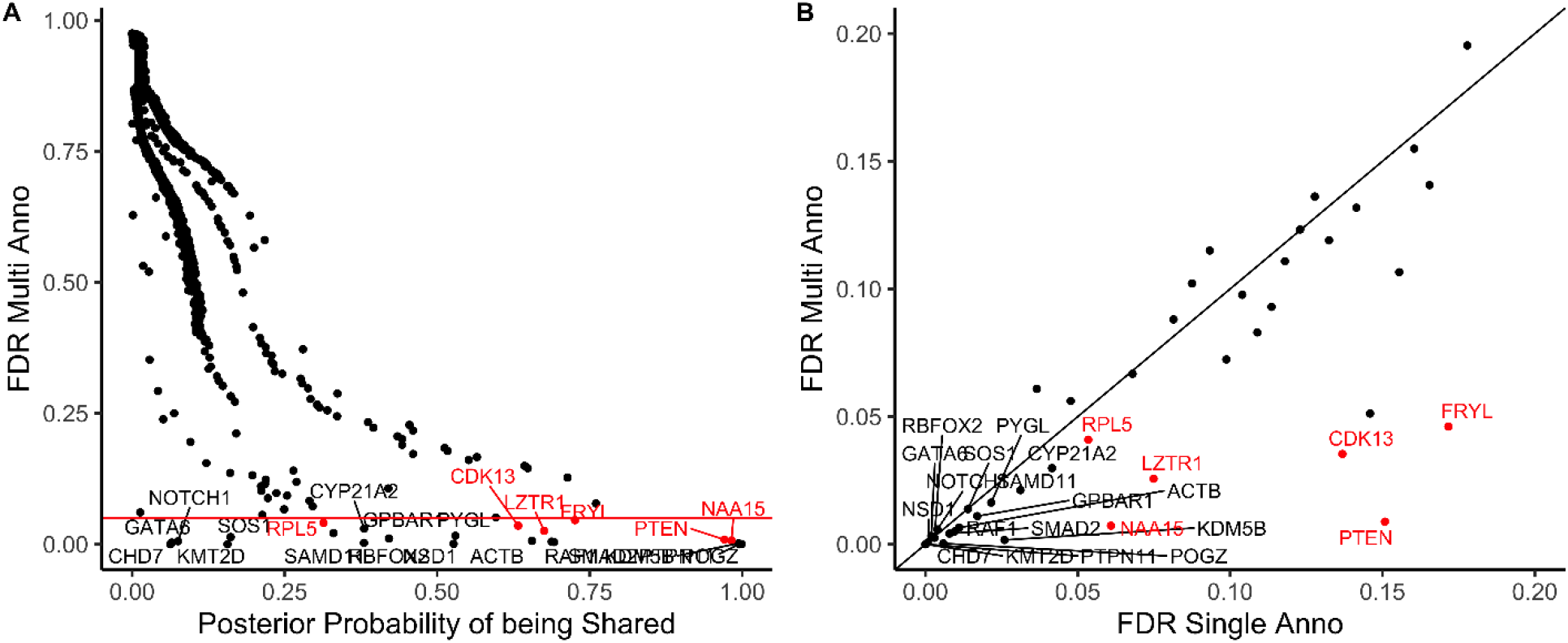
Multi-trait analyses prioritized additional genes with high posterior probability of being shared risk genes for CHD. Gene names of the 23 genes identified by the multi-trait model with annotations are shown on the plot and the additional 6 genes that were identified by the multi-trait models are marked in red. (A) shows that the 6 additional genes identified by the multi-trait models had high posterior probability of being shared. The x-axis represents the posterior probability of being shared calculated from the multi-trait model with annotations. The y-axis represents the FDR of genes calculated from the multi-trait model with annotations. (B) shows that the top genes in the multi-trait model with annotations also had low FDR (<0.2) in the single-trait model with annotations. The x-axis represents the FDR of genes calculated from the single-trait model with annotations. The y-axis represents the FDR of genes calculated from the multi-trait model with annotations.

**Fig 7.**
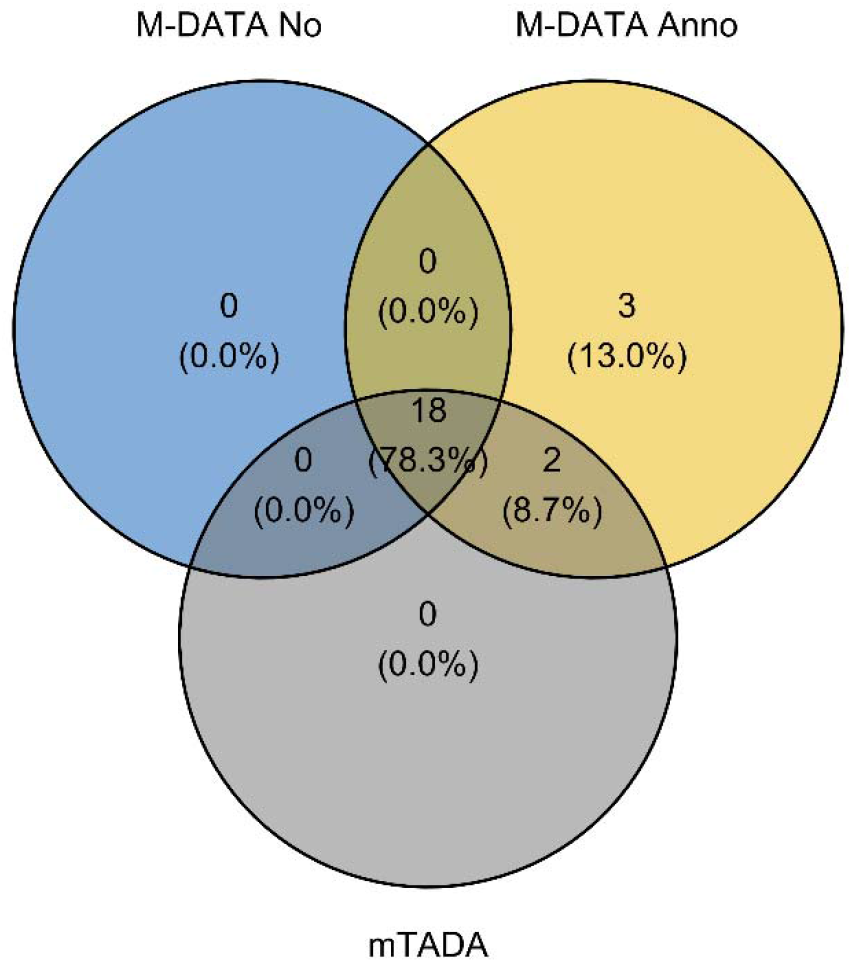
Venn diagram of genes identified by M-DATA and mTADA for CHD. M-DATA multi-trait model with annotations identified 3 additional genes (*CDK13, SAMD11* and *RPL5*).

We take the 5 CHD genes identified by the multi-trait models, but not the single-trait models as examples to demonstrate the pleiotropic effect. We selected the DNM counts of CHD and autism, FDR of the single-trait model with annotations and FDR of the multi-trait model with annotations model from the results (Table 2). From this table, we can see *CDK13, FRYL, LZTR1, NAA15* and *PTEN* have 2 DNM counts for CHD and at least 1 shared DNM count with autism. For *PTEN*, it has 4 shared counts with autism, and we can see a substantial increase of significance in terms of FDR. Thus, the insight is that genes with shared counts with autism are more likely to be prioritized for CHD in multi-trait analyses by leveraging the pleiotropic effect.

**Table 2.** Pleiotropic effect boosts power for M-DATA multi-trait models.

| Gene | CHD Counts | Autism Counts | FDR Single Anno | FDR Multi Anno |
| --- | --- | --- | --- | --- |
| *CDK13* | 2 | 1 | 0.137 | 0.0353 |
| *FRYL* | 2 | 2 | 0.172 | 0.0461 |
| *LZTR1* | 2 | 1 | 0.0749 | 0.0257 |
| *NAA15* | 2 | 3 | 0.0609 | 0.00726 |
| *PTEN* | 2 | 4 | 0.151 | 0.00882 |

**Table 3.** Comparison of M-DATA multi-trait models with mTADA.

|  | M-DATA No | M-DATA Anno | mTADA |
| --- | --- | --- | --- |
| CHD | 18 | 23 | 20 |
| Autism | 28 | 37 | 28 |

Among the 23 identified genes from joint model with annotations, 11 were well established known CHD genes based on a previously compiled gene list with 254 known CHD genes [4]. They are involved in essential developmental pathways or biological processes, such as Notch signaling (*NOTCH1*), RAS signaling (*PTPN11, RAF1, SOS1*), PI3K/AKT signaling (*PTEN*), chromatin modeling (*CHD7, KMT2D, NSD1*), transcriptional regulations (*GATA6*), and cell structural support (*ACTB, RPL5*) [42, 43].

Among the 12 novel genes, *RBFOX2, SMAD2, CDK13* are three emerging CHD risk genes that have been recently reported to cause hypoplastic left heart syndrome [9, 44, 45], laterality defect [1, 46], and septal defects and pulmonary valve abnormalities [47], respectively.

Additionally, 4 novel genes, *POGZ, KDM5B, NAA15*, and *FRYL*, harbored at least two *de novo* mutations in both CHD and autism cohorts.

*POGZ*, encoding a heterochromatin protein 1 alpha-binding protein, participates in chromatin modeling and gene regulations. It binds to chromatin and facilitates the packaging of DNA onto chromosomes. *POGZ* damaging *de novo* mutations were strongly linked with autism spectrum disorders and other neurodevelopmental disorders [48, 49]. Interestingly, one of the reported mutation carriers also presented cardiac defect [50].

*KDM5B* is a lysine-specific histone demethylase. Studies have shown that it regulates H3K4 methylation near promoter and enhancer regions in embryonic stem cells and controls the cell pluripotency [51, 52]. The deletion of *KDM5B* in mice is neonatal lethal with respiratory failure and neurodevelopmental defects [53]. Recessive mutations in the gene were associated with mental retardation (OMIM: 618109) and one reported patient presented atrial septal defect.

*NAA15* encodes the auxiliary subunit of N-Alpha-Acetyltransferase 15, which catalyzes one of the most common post-translational modification essential for normal cell functions. Protein-truncating mutations in *NAA15* were reported in intellectual disability and autism patients, some of whom also presented a variety of cardiac abnormalities including ventricular septal defect, heterotaxy, pulmonary stenosis and tetralogy of Fallot [54].

*POGZ, KDM5B*, and *NAA15* are all highly expressed in developmental heart at mice embryonic day E14.5 [4]. *POGZ* and *NAA15* are intolerant for both LoF and missense mutations, given that they have a pLI score > 0.9 and a missense z-score > 3. *KDM5B* is intolerant for missense mutations with a missense z-score of 1.78. Considering their intolerance of protein-altering variants, the identification of damaging *de novo* mutations in them is highly unlikely. Therefore, our analyses suggest that *POGZ, KDM5B* and *NAA15* may be considered as new candidate CHD genes.

Furthermore, among the 17 genes with at least one *de novo* mutation in CHD and autism cohorts, 5 genes (*KMT2D, NSD1, POGZ, SMAD2, KDM5B*) play a role in chromatin modeling. Such high proportion is consistent with previous studies that chromatin modeling-related transcriptional regulations are essential for both cardiac and neuro-development, and genes with critical regulatory roles in the process may be pleotropic [9].

Further, we compared the performance of M-DATA with mTADA [23] using the same real data of CHD and autism. We fitted both methods with damaging mutations (LoF and Dmis mutations). mTADA identified all 18 genes identified by our no annotation model, and missed 3 genes (*CDK13, SAMD11*, and *RPL5*) identified by our annotation model for CHD (Table 2). We visualized the results with Venn diagrams (Fig 6 and Fig J in the S1 Text). We also compared our results with the results of CHD-ASD pair reported by mTADA using CHD data [55] autism data [11], and mutability data downloaded from the github webpage of mTADA (Table E in the S1 Text).

## Discussion

In this paper, we have introduced M-DATA, a method to jointly analyze *de novo* mutations from multiple traits by integrating shared genetic information across traits. The implemented model is available at https://github.com/JustinaXie/MDATA. This approach can increase the effective sample size for all traits, especially for those with small sample size. M-DATA also provides a flexible framework to incorporate external functional annotations, either variant-level or gene-level, which can further improve the statistical power. Through simulation study, we demonstrated that our multi-trait model with annotations could not only gain accurate estimates on the proportion of shared risk genes between two traits and the proportion of risk genes for a single trait under various settings, but also gained statistical power compared to the single-trait models. In addition, M-DATA adopts the Expectation-Maximization (EM) algorithm in estimation, which does not require prior parameter specification or pre-estimation. In our simulation study, we found that the algorithm converges faster than methods that use MCMC for estimation (Table D in the S1 Text).

Despite the success, there are some limitations in the current M-DATA model. In our real data analysis, we used two different data sources for CHD and autism. Samples with both diseases in our multi-trait analysis may bring bias because of the violation of independence assumption in our multi-trait models. The autism DNM data in our analysis are from different studies, and different filtering criteria across studies may also bring bias and dilute our signals. In addition, we only considered two traits simultaneously. Though it is straightforward to extend our model to more than two traits, the number of groups (i.e., the dimension of latent variables *Z*_*i*_) increases exponentially with the number of traits (2^*N*^ for N traits) [23]. This might bring difficulty in estimation and have more computational cost. Model performance with more than two traits need further exploration. Currently, we did not consider the influence of admixed population in M-DATA. In a recent study, Kessler et al. studied DNM across 1,465 diverse genomes and discovered mutation rates may be affected by the environment more significantly than previously known [56]. Confounding from the environment on mutation rates could be further explored through cross-ancestry rare variant studies.

In conclusion, M-DATA is a novel and powerful approach to performing gene-based association analysis for DNMs across multiple traits. Not only does M-DATA have better statistical power than single-trait methods, it also provides reasonable estimation of shared proportion of risk genes between two traits, which gives novel insights in the understanding of disease mechanism. We have successfully applied M-DATA to study CHD, which identified significant 23 genes for our multi-trait model with annotations. Moreover, our method provides a general framework in extending single-trait method to multi-trait method which can also incorporate information from functional annotations. Recently, there are several advancements in the association analysis for rare variants, such as jointly analyzing DNMs and transmitted variants [41], analyzing DNMs from whole-genome sequencing (WGS) data [25], and incorporating pathway information [57]. Extension of these methods to multi-trait analysis is a potential future direction.

## Data Availability

CHD data were downloaded from the supplement of Jin et al (PMID: 28991257). Autism data were acquired from denovo-db.

https://pubmed.ncbi.nlm.nih.gov/28991257/

https://denovo-db.gs.washington.edu/denovo-db/Usage.jsp

## Ethics Statement

This study is approved by Yale Human Research Protection Program Institutional Review Boards (IRB protocol ID 2000028735).

## Supporting Information

**S1 Table. Simulation of Estimation and Power Evaulation.**

**S2 Table. Simulation of Comparison with mTADA**.

**S3 Table. Results of Real Data Application**.

**S1 Text. Supplementary Notes on Methods and Results**.

## Acknowledgements

Supported in part by NIH grant R03HD100883-01A1. We thank Dr. Sheng Chih (Peter) Jin, Geyu Zhou and Hanmin Guo for helpful discussions.

