## Supplemental Text 1 for "M-DATA: A Statistical Approach to Jointly Analyzing *De Novo* Mutations for Multiple Traits"

**Supporting Information**

### **Details of Functional Annotation and Feature Selection**

For variant-level annotations, we first benchmarked deleterious missense variants by MetaSVM (PolyPhen(P), PolyPhen(D), MPC, CADD, and REVEL scores were set as 0 if the variant was not predicted as Dmis by MetaSVM). For variants not existing in the gnomAD database, we set the value of variant as 1/(125748*2) for gnomAD_exome allele frequency and 1/(71702*2) for gnomAD_genome allele frequency. We took the -log10 transformation for both allele frequencies as our variant-level allele frequency scores. Then, we filled all NAs with 0s. After these steps, all variant-level annotations were preprocessed as described in Table A(a). An example of preprocessing steps for gene *ATCB* is provided in Table A(c). For gene-level annotations, they can be directly used as gene-level features (Table A(b)). We normalized all continuous gene-level features before model fitting.

For single-trait analyses, we first checked the correlation of gene-level features. We detected highly correlated clusters (pairwise Pearson’s correlations >0.7) for both CHD and autism. For CHD, the clusters are MPC, CADD and REVEL, dbsc_SNV_ADA_SCORE and dbscSNV_RF_SCORE, and LoF and dpsi_zscore. We kept CADD, dbscSNV_RF_SCORE, and LoF in these clusters and removed other correlated annotations. For autism, the clusters are REVEL and CADD, dbsc_SNV_ADA_SCORE and dbscSNV_RF_SCORE, and gnomAD_exome_ALL and gnomAD_genome_ALL. To be consistent with CHD, we kept CADD and dbscSNV_RF_SCORE in the first two clusters. We selected gnomAD_exome_ALL from the third cluster. We conducted single-trait analyses for CHD and autism with the selected annotations. We removed features with weak effect sizes (absolute values $\leq$0.01) and refit the model. For multi-trait analyses, we constructed the annotation matrices using the features selected from single-trait analyses.

(a) **Variant-level annotations**

| **Annotation** | **ANNOVAR varid** | **Classification** | **Preprocessing** |
| --- | --- | --- | --- |
| Polyphen (D) | Polyphen2_HVAR_pred | Deleterious (Categorical) | D->1 |
| Polyphen (P) | Polyphen2_HVAR_pred | Deleterious (Categorical) | P->1 |
| MPC | MPC | Deleterious (Score 0-5) | Normalize |
| CADD | CADD13_PHRED | Deleterious (Score 0-100) | Normalize |
| REVEL | REVEL | Deleterious  (Score 0-1) | Normalize |
| LoF | ExonicFunc.refGene  Func.refGene | Deleterious (Categorical) | Frameshift insertion/ deletion, stopgain, stoploss, splicing site alternation -> 1 |
| gnomAD_exome | gnomAD_exome_ALL | Allele Frequency (Frequency) | Normalize |
| gnomAD_genome | gnomAD_genome_ALL | Allele Frequency (Frequency) | Normalize |
| dbscSNV_ADA_score | dbscSNV_ADA_SCORE | Splicing (Score 0-1) | Binarize:  dbscSNV_ADA_SCORE >0.6 |
| dbscSNV_RF_score | dbscSNV_RF_SCORE | Splicing (Score 0-1) | Binarize:  dbscSNV_RF_SCORE >0.6 |
| dpsi_zscore | dpsi_zscore | Splicing  (z-score) | Binarize:  \|dpsi_zscore \|>2 |

(b) **Gene-level annotations**

| **Annotation** | **Source** | **Classification** |
| --- | --- | --- |
| pLI | gnomAD | Conservation (0-1) |
| mis_z | gnomAD | Conservation (0-1) |

**(c) An example of collapsing variant-level annotations for gene *ACTB***

|  | **PolyPhen (P)** | **……** | **CADD** | **dpsi_zscore** |
| --- | --- | --- | --- | --- |
| **Mutation A1** | B -> 0 |  | 23.6 -> 1.56 | -0.009 -> 0 |
| **Mutation A2** | P -> 1 |  | 24.8 -> 3.20 | 0.842 -> 0 |
| **Mutation A3** | NA -> 0 |  | (NA -> 0)->-1.07 | NA -> 0 |
| **Gene *ATCB*** | 0+1+0=1 |  | 1.56+3.20-1.07=3.69 | 0 |

**Table A. Summary for Functional Annotations.** For a vector $x$, normalization is defined as $\frac{x-\bar{x}}{\mathrm{sd}(x)}$, where  $\bar{x}$ is the mean of $x$ and $\mathrm{sd}(x)$ is the standard deviation of $x$. (a) Summary of variant-level annotations from the output of ANNOVAR [1]. (b) Summary of gene-level annotations retrieved from gnomAD [2]. (c) An example of collapsing variant-level annotations to gene-level scores for gene *ACTB.*

**(a)**

| **Gene** | **FDR** | **Shared Counts** | **Annotation** |
| --- | --- | --- | --- |
| *CHD7* | 6.55E-17 | 1 | H-CHD |
| *KMT2D* | 1.21E-15 | 1 | H-CHD |
| *PTPN11* | 4.32E-15 | 3 | H-CHD |
| *POGZ* | 0.000306 | 3 |  |
| *NSD1* | 0.000774 | 2 | H-CHD |
| *KDM5B* | 0.00167 | 3 |  |
| *RBFOX2* | 0.00254 | 0 |  |
| *GATA6* | 0.00325 | 0 | H-CHD |
| *SMAD2* | 0.00417 | 1 |  |
| *RAF1* | 0.005 | 1 | H-CHD |
| *NOTCH1* | 0.00581 | 2 | H-CHD |
| *ACTB* | 0.00649 | 1 | H-CHD |
| *NAA15* | 0.00726 | 2 |  |
| *PTEN* | 0.00882 | 2 | H-CHD |
| *GPBAR1* | 0.011 | 0 |  |
| *SOS1* | 0.0138 | 0 | H-CHD |
| *PYGL* | 0.0163 | 1 |  |
| *SAMD11* | 0.0212 | 0 |  |
| *LZTR1* | 0.0257 | 1 |  |
| *CYP21A2* | 0.0298 | 0 |  |
| *CDK13* | 0.0354 | 1 |  |
| *RPL5* | 0.0409 | 0 | H-CHD |
| *FRYL* | 0.0461 | 2 |  |

**(b)**

| **Gene** | **FDR** | **Shared Counts** | **Satterstrom et al. [3]** |
| --- | --- | --- | --- |
| *SCN2A* | 1.83E-14 | 0 | Yes |
| *ARID1B* | 4.47E-11 | 1 | Yes |
| *KDM5B* | 1.55E-07 | 3 | Yes |
| *POGZ* | 9.14E-07 | 3 | Yes |
| *SHANK3* | 6.53E-06 | 0 | Yes |
| *KMT5B* | 1.16E-05 | 0 | No |
| *SYNGAP1* | 2.83E-05 | 0 | Yes |
| *PTEN* | 6.09E-05 | 2 | Yes |
| *SLC6A1* | 8.63E-05 | 0 | Yes |
| *WDFY3* | 0.000109 | 0 | No |
| *ADNP* | 0.00017 | 0 | Yes |
| *NAA15* | 0.000277 | 2 | No |
| *ANKRD11* | 0.000433 | 0 | Yes |
| *DNMT3A* | 0.000598 | 0 | Yes |
| *KDM6B* | 0.000758 | 1 | Yes |
| *PTPN11* | 0.00102 | 3 | No |
| *ASH1L* | 0.00146 | 1 | Yes |
| *ANK2* | 0.00203 | 1 | Yes |
| *ASXL3* | 0.00308 | 0 | Yes |
| *HIVEP3* | 0.00424 | 0 | No |
| *PHF3* | 0.0056 | 0 | No |
| *CHD8* | 0.00743 | 0 | Yes |
| *SET* | 0.00941 | 0 | No |
| *KIRREL3* | 0.0114 | 0 | No |
| *WAC* | 0.0134 | 1 | Yes |
| *SPAST* | 0.0155 | 0 | Yes |
| *KMT2C* | 0.0177 | 2 | Yes |
| *FOXG1* | 0.02 | 0 | No |
| *FOXP1* | 0.023 | 0 | Yes |
| *LMTK3* | 0.0257 | 0 | No |
| *BRF1* | 0.0287 | 1 | No |
| *UNC5B* | 0.0316 | 1 | No |
| *PRKAR1B* | 0.0346 | 0 | No |
| *GIGYF1* | 0.0375 | 1 | Yes |
| *NUDT17* | 0.0405 | 0 | No |
| *HECTD4* | 0.0437 | 0 | Yes |
| *UBN2* | 0.048 | 0 | No |

**Table B. Significant genes (FDR<0.05) identified by M-DATA.** M-DATA was performed on 2,645 CHD probands and 5,623 autism probands. The column *Shared Count* is the minimum number of DNMs in the CHD and autism datasets for the significant genes. (a) The 23 significant genes (FDR<0.05) identified by M-DATA multi-trait model with annotations for CHD. The last column indicates whether they were known human CHD genes (H-CHD) [4]. (b) The 37 significant genes (FDR<0.05) identified by M-DATA multi-trait model with annotations for autism. The last column indicates whether they were significant genes in Satterstrom et al. [3]

| **Parameter** | **Value** |
| --- | --- |
| $\hat{\pi}_{00}$ | 0.9410 |
| $\hat{\pi}_{10}$ | 0.0135 |
| $\hat{\pi}_{01}$ | 0.0343 |
| $\hat{\pi}_{11}$ | 0.0116 |

|  | **Risk Gene Proportion** |
| --- | --- |
| CHD | $\hat{\pi}_{10}+\hat{\pi}_{11}=$ 0.0251 |
| Autism | $\hat{\pi}_{01}+\hat{\pi}_{11}=$ 0.0459 |

**Table C. Parameter estimates of M-DATA multi-trait analysis of CHD and autism.** The estimated risk gene proportions correspond to the hypotheses in Methods. The estimated values of risk gene proportion for CHD and autism were calculated by adding up the proportion of corresponding hypotheses.

| **Setting** | | | **Time (minutes)** | |
| --- | --- | --- | --- | --- |
| $N$ | $\pi_{11}$ | $\beta$ | M-DATA | mTADA |
| 2000 | 0.01 | 0.1 | 0.13 | 26.45 |
| 2000 | 0.01 | 0.3 | 0.11 | 25.59 |
| 2000 | 0.01 | 0.5 | 0.11 | 25.08 |
| 2000 | 0.03 | 0.1 | 0.13 | 24.94 |
| 2000 | 0.03 | 0.3 | 0.11 | 24.47 |
| 2000 | 0.03 | 0.5 | 0.12 | 24.55 |
| 2000 | 0.05 | 0.1 | 0.13 | 24.68 |
| 2000 | 0.05 | 0.3 | 0.11 | 24.58 |
| 2000 | 0.05 | 0.5 | 0.12 | 24.8 |
| 2000 | 0.07 | 0.1 | 0.13 | 25.94 |
| 2000 | 0.07 | 0.3 | 0.11 | 25.06 |
| 2000 | 0.07 | 0.5 | 0.11 | 25.02 |
| 2000 | 0.09 | 0.1 | 0.12 | 30.93 |
| 2000 | 0.09 | 0.3 | 0.11 | 29.32 |
| 2000 | 0.09 | 0.5 | 0.12 | 26.22 |
| 5000 | 0.01 | 0.1 | 0.08 | 25.47 |
| 5000 | 0.01 | 0.3 | 0.09 | 25.21 |
| 5000 | 0.01 | 0.5 | 0.11 | 25.34 |
| 5000 | 0.03 | 0.1 | 0.09 | 24.7 |
| 5000 | 0.03 | 0.3 | 0.09 | 24.64 |
| 5000 | 0.03 | 0.5 | 0.11 | 24.67 |
| 5000 | 0.05 | 0.1 | 0.08 | 24.61 |
| 5000 | 0.05 | 0.3 | 0.09 | 24.79 |
| 5000 | 0.05 | 0.5 | 0.1 | 24.82 |
| 5000 | 0.07 | 0.1 | 0.09 | 24.91 |
| 5000 | 0.07 | 0.3 | 0.09 | 24.79 |
| 5000 | 0.07 | 0.5 | 0.1 | 25.06 |
| 5000 | 0.09 | 0.1 | 0.09 | 26.09 |
| 5000 | 0.09 | 0.3 | 0.09 | 25.33 |
| 5000 | 0.09 | 0.5 | 0.1 | 25.32 |
| 10000 | 0.01 | 0.1 | 0.08 | 25.15 |
| 10000 | 0.01 | 0.3 | 0.08 | 24.93 |
| 10000 | 0.01 | 0.5 | 0.1 | 25.26 |
| 10000 | 0.03 | 0.1 | 0.07 | 24.76 |
| 10000 | 0.03 | 0.3 | 0.08 | 24.87 |
| 10000 | 0.03 | 0.5 | 0.1 | 24.83 |
| 10000 | 0.05 | 0.1 | 0.08 | 24.83 |
| 10000 | 0.05 | 0.3 | 0.08 | 24.78 |
| 10000 | 0.05 | 0.5 | 0.1 | 24.83 |
| 10000 | 0.07 | 0.1 | 0.08 | 24.8 |
| 10000 | 0.07 | 0.3 | 0.08 | 25.23 |
| 10000 | 0.07 | 0.5 | 0.1 | 24.84 |
| 10000 | 0.09 | 0.1 | 0.08 | 25.39 |
| 10000 | 0.09 | 0.3 | 0.08 | 25.67 |
| 10000 | 0.09 | 0.5 | 0.1 | 25.27 |

**Table D. Time comparison of M-DATA and mTADA under multiple settings in the simulation study.** The computational time was benchmarked on an Intel Xeon Gold 6240 processor (2.60 GHZ).

### **Comparison with the Results Reported in mTADA [5]**

We fitted our multi-trait models by plugging in the sum of LoF and Dmis mutation count of a gene as the mutation count for a gene in our model and the sum of LoF and Dmis mutability of a gene as the mutability for our model. This model is defined as M-DATA multi-trait model without annotation in the comparison. For M-DATA multi-trait model with annotations, we used the LoF mutation count of each gene as the annotation. Following the risk gene identification method in mTADA, we defined genes with posterior probability (PP) larger than 0.8 as risk genes. For CHD data, among 19,358 genes tested, mTADA identified 11 risk genes, with 9 of 11 are unknown human CHD genes, M-DATA multi-trait model without annotation identified 17 risk genes, with 13 unknown human CHD genes, and M-DATA multi-trait model with annotations identified 19 genes, with 15 unknown human CHD genes. All 11 risk genes identified by mTADA were also identified by our two models. The 6 additional unknown human CHD genes identified by our models are *BRD4, CPD, GANAB, NCKAP1, RABGAP1L* and *SMAD2*. If we relax the threshold of mTADA to PP>0.5, all 6 genes can also be found by mTADA (Table E).

| **Number of Genes** | **M-DATA**  **No Anno** | **M-DATA**  **Anno** | **mTADA** |
| --- | --- | --- | --- |
| M-DATA CHD | 17 | 19 | 11 |
| Known Human CHD [4] | 4 | 4 | 2 |
| Overlap CHD with mTADA | 11/11 | 11/11 | - |
| M-DATA Autism | 33 | 75 | 45 |
| Overlap Autism with Satterstrom et al. [3] | 25 | 40 | 27 |
| Overlap Autism with mTADA | 30/45 | 34/45 | - |

**Table E. Comparison of M-DATA multi-trait models with the results reported in the mTADA paper [5].**

(a) **One annotation is effective**

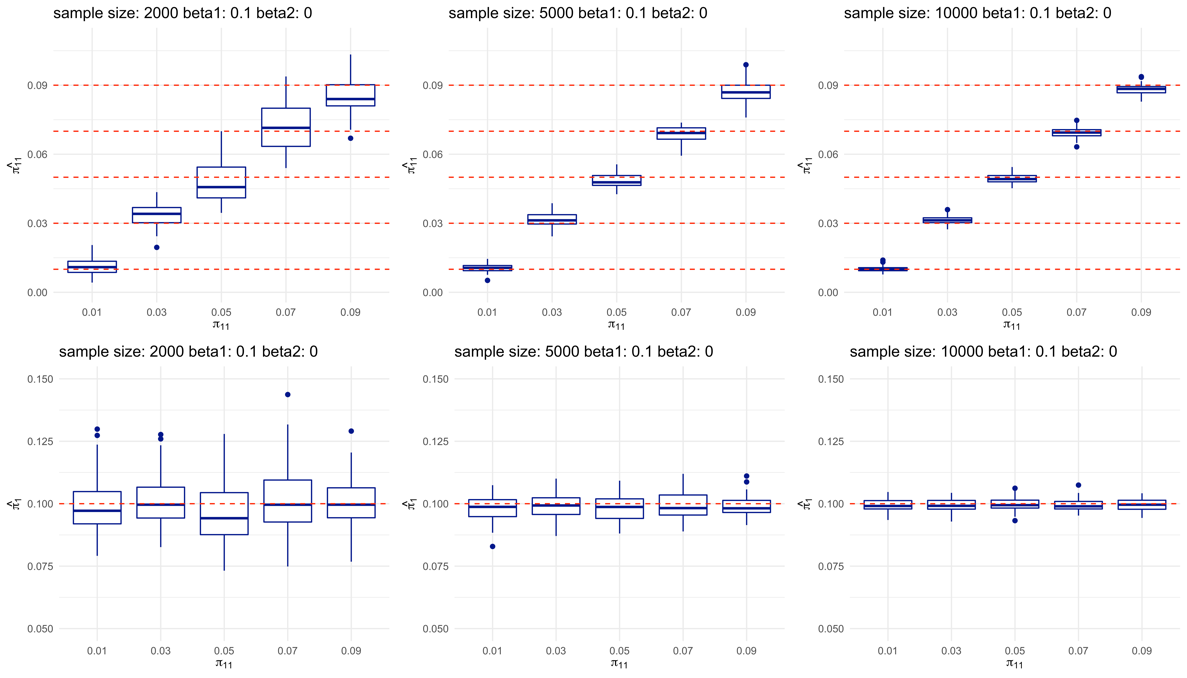

(b) **No annotation is effective**

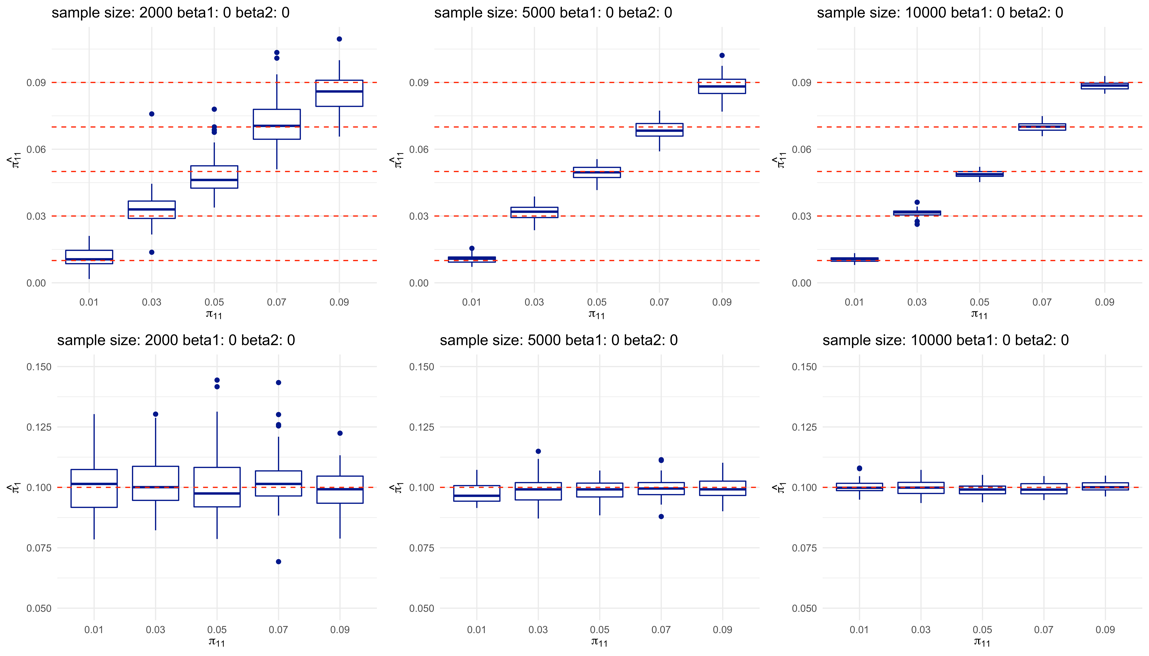

Fig A. Multi-trait analysis can accurately estimate the proportion of shared risk genes and single-trait risk genes when one annotation is effective or no annotation is effective. For both (a) and (b), top panels show the estimation of shared risk proportion ($\hat{\pi}_{11})$, and bottom panels show the estimation of risk gene proportion for a single trait ${(\hat{\pi}}_{11}+\hat{\pi}_{10})$. For each panel, each plot from left to right represents study sample size of 2000, 5000, and 10000, respectively. Within each plot, boxes from left to right represent the proportion of shared risk genes $\pi_{11}$being 0.01, 0.03, 0.05, 0.07 and 0.09, respectively. Each scenario is replicated for 50 times in our simulations. True values are shown in dashed red lines.

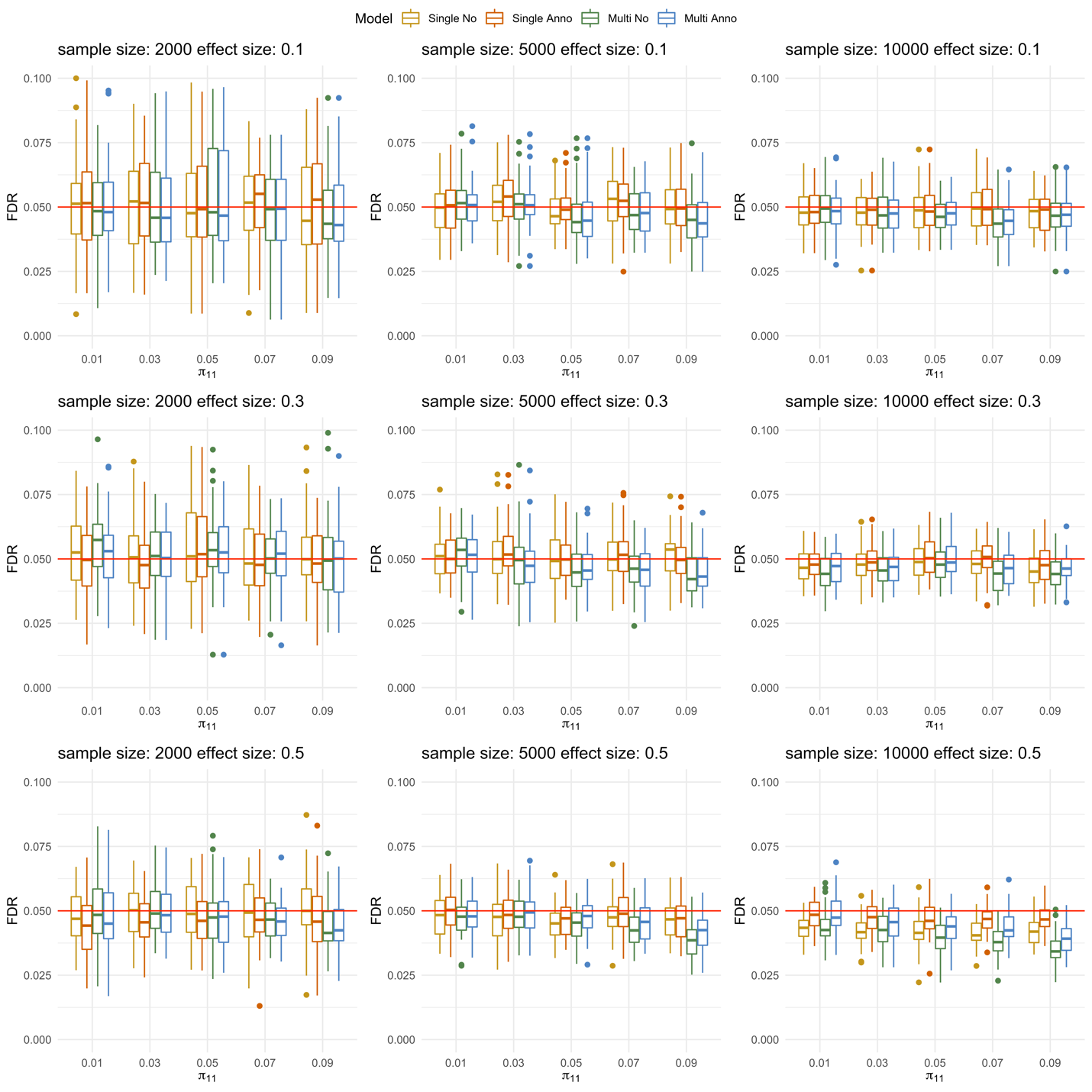

Fig B. M-DATA can control FDR under different simulation settings. Panels from top to bottom show the powers under weak, moderate and strong annotations, respectively. For each panel, each plot from left to right represents study sample size of 2000, 5000, and 10000, respectively. Within each plot, boxes from left to right represent the proportion of shared risk genes $(\pi_{11})$ being 0.01, 0.03, 0.05, 0.07 and 0.09, respectively. Each scenario is replicated for 50 times in our simulations. The expected value of FDR (0.05) is shown in red lines.

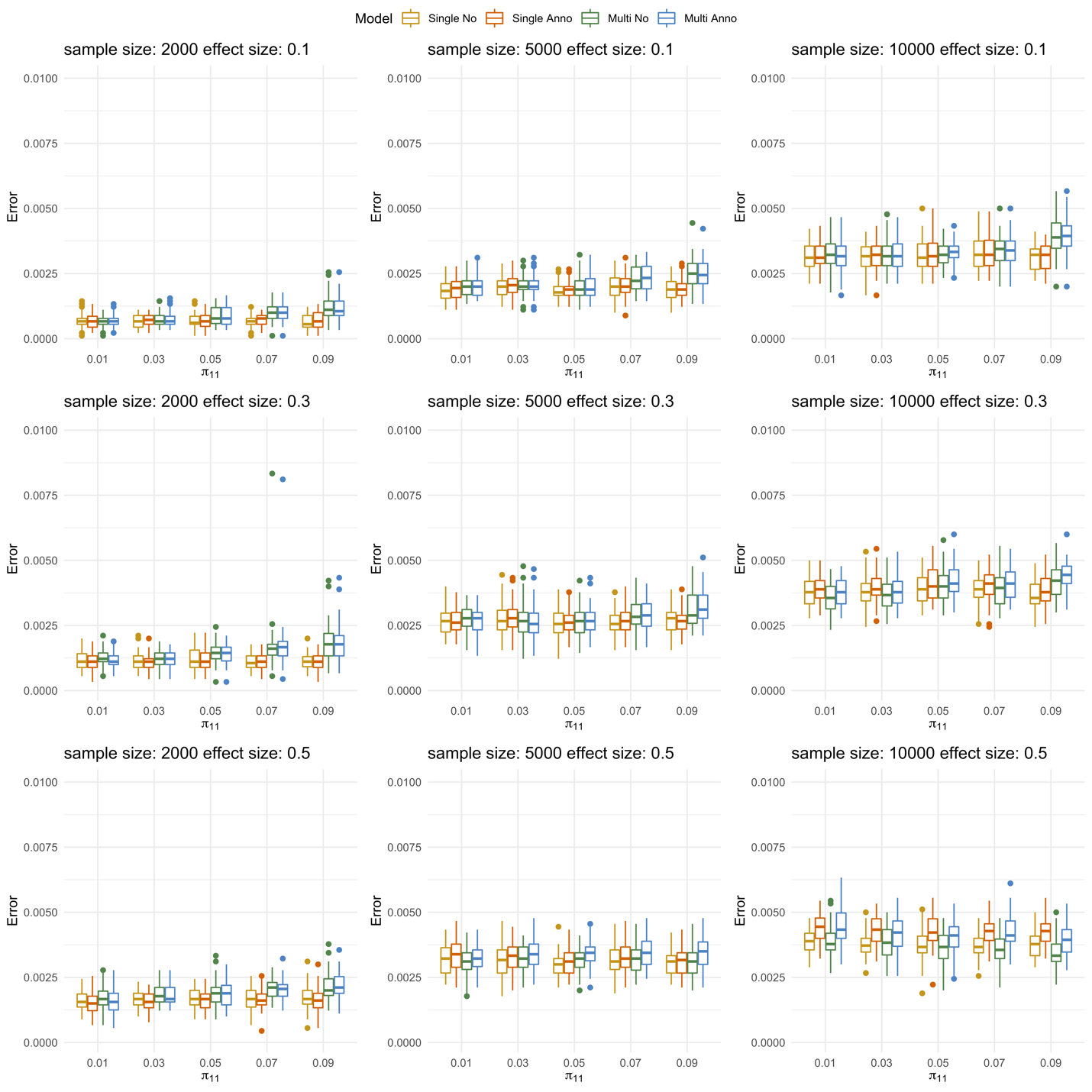

Fig C. Type 1 errors under different simulation settings. Panels from top to bottom show the type 1 errors under weak, moderate and strong annotations, respectively. For each panel, each plot from left to right represents study sample size of 2000, 5000, and 10000, respectively. Within each plot, boxes from left to right represent the proportion of shared risk genes ($\pi_{11})$ being 0.01, 0.03, 0.05, 0.07 and 0.09, respectively. Each scenario is replicated for 50 times in our simulations.

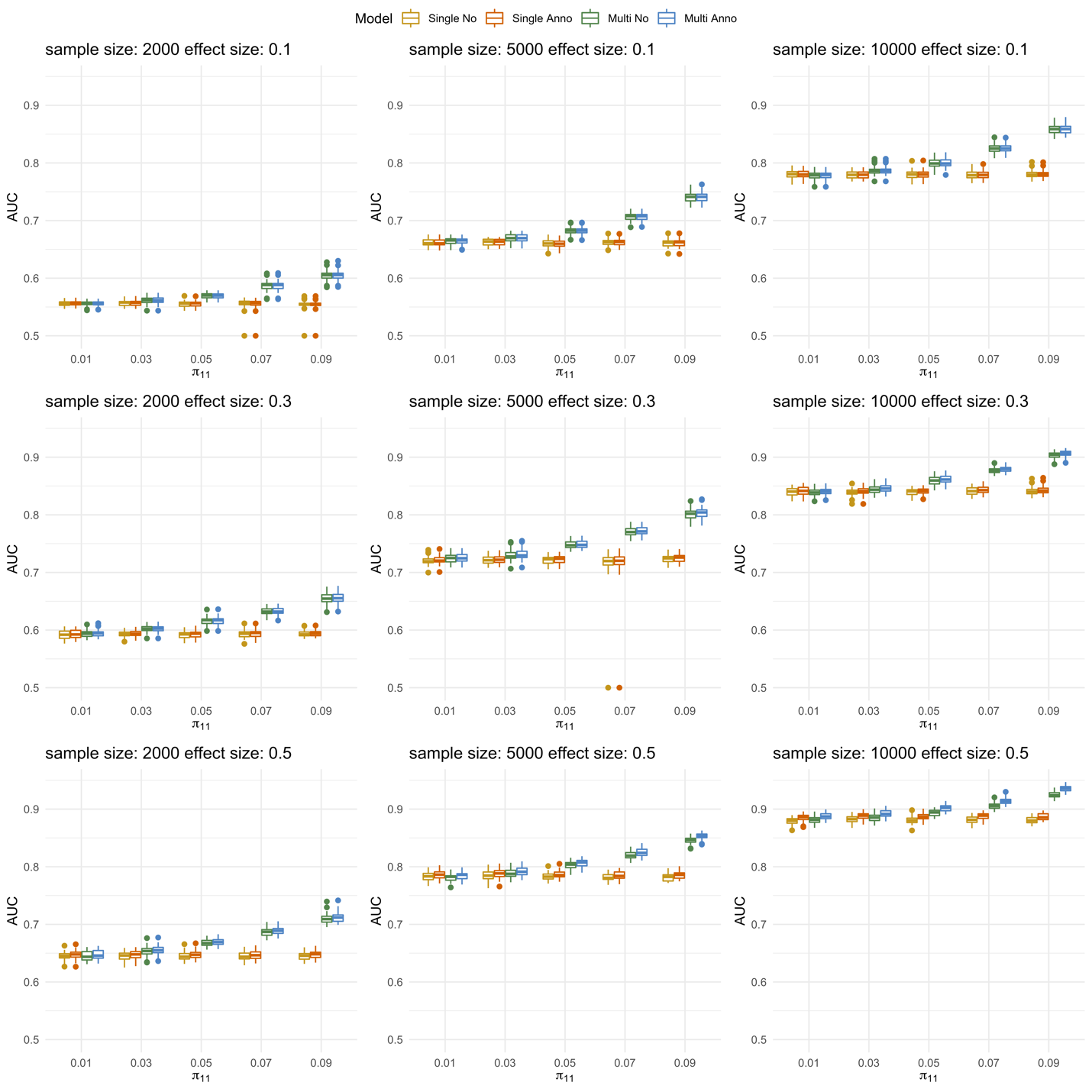

Fig D. AUCs under different simulation settings. Panels from top to bottom show the AUCs under weak, moderate and strong annotations. For each panel, each plot from left to right represents study sample size of 2000, 5000, and 10000, respectively. Within each plot, boxes from left to right represent the proportion of shared risk genes ($\boldsymbol{\pi}_{\boldsymbol{11}}\boldsymbol{)}$ being 0.01, 0.03, 0.05, 0.07 and 0.09, respectively. Each scenario is replicated for 50 times in our simulations.

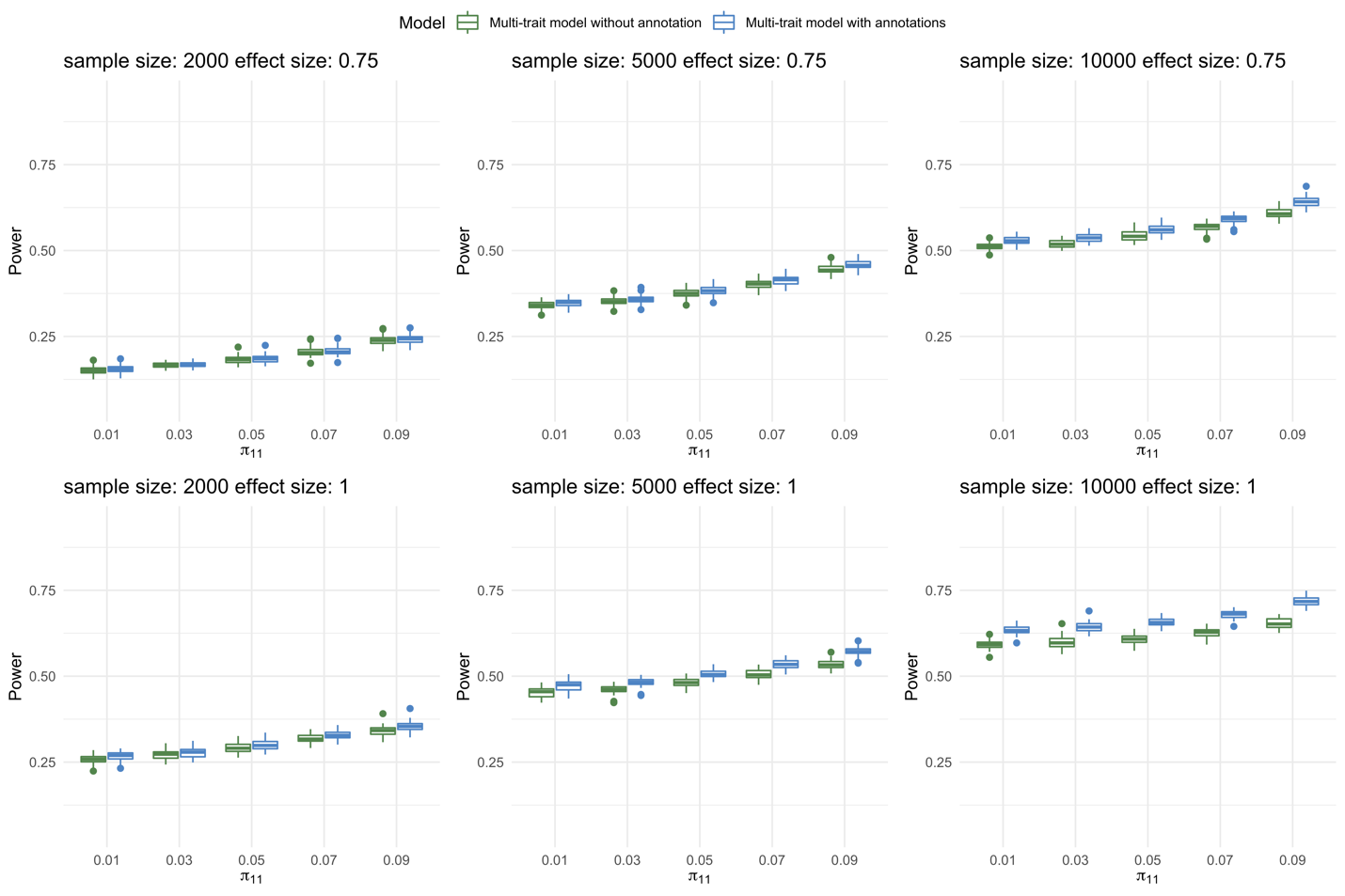

Fig E. Multi-trait model with annotations has more increase in power than multi-trait model without annotation when the effect size of annotations is stronger. The top and bottom panels present the cases when the effect size of annotations ${(\beta_{j0},\beta}_{j1},\beta_{j2}, \beta_{j3}), j=1,2$are (2,0.75,0.75,0.75) and (2,1,1,1), respectively. For each panel, each plot from left to right represents study sample size of 2000, 5000, and 10000, respectively. Within each plot, boxes from left to right represent the proportion of shared risk genes ($\boldsymbol{\pi}_{\boldsymbol{11}}$) being 0.01, 0.03, 0.05, 0.07 and 0.09, respectively. Each scenario is replicated for 50 times in our simulations.

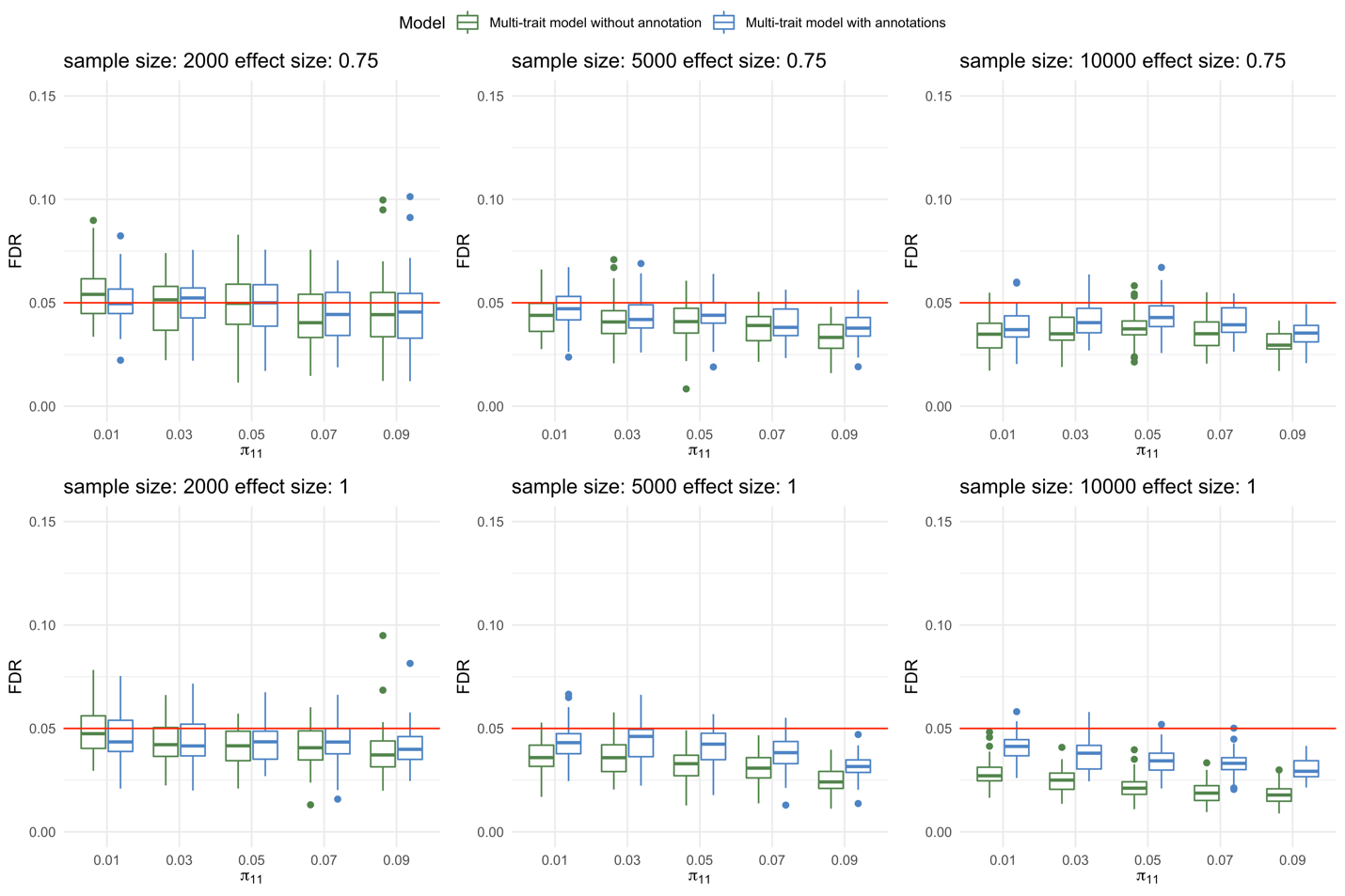

Fig F. Multi-trait models can control FDR when the effect size of annotations is stronger. The top and bottom panels present the cases when the effect size of annotations ${\mathbf{(}\boldsymbol{\beta}_{\boldsymbol{j}\mathbf{0}}\mathbf{,}\boldsymbol{\beta}}_{\boldsymbol{j}\mathbf{1}}\mathbf{,}\boldsymbol{\beta}_{\boldsymbol{j}\mathbf{2}}\mathbf{,}\boldsymbol{\beta}_{\boldsymbol{j}\mathbf{3}}\mathbf{),}\boldsymbol{j}\mathbf{=1,2}$are (2,0.75,0.75,0.75) and (2,1,1,1), respectively. For each panel, each plot from left to right represents study sample size of 2000, 5000, and 10000, respectively. Within each plot, boxes from left to right represent the proportion of shared risk genes ($\boldsymbol{\pi}_{\boldsymbol{11}}$) being 0.01, 0.03, 0.05, 0.07 and 0.09, respectively. Each scenario is replicated for 50 times in our simulations. The expected value of FDR (0.05) is shown in red lines.

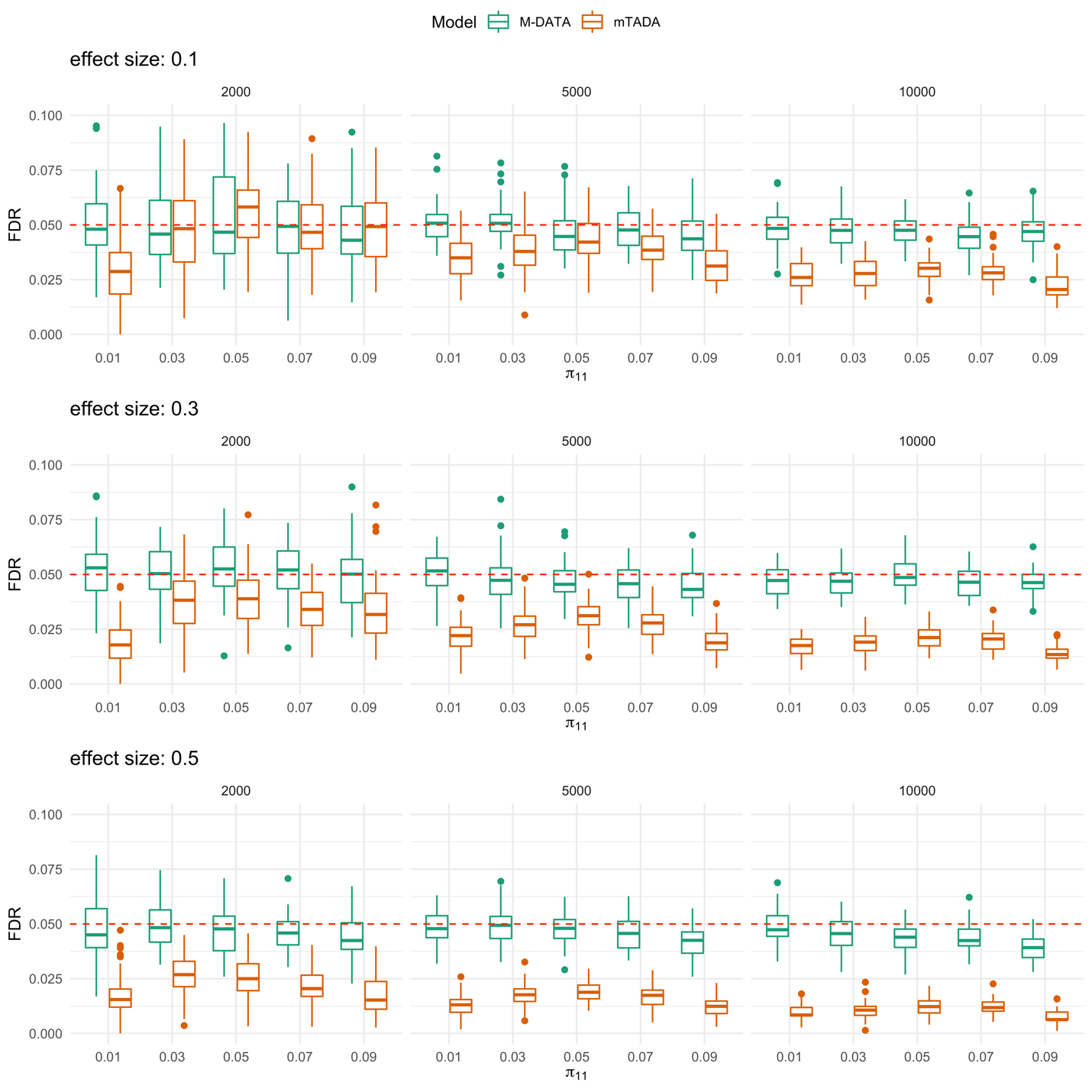

Fig G. mTADA is more conservative than M-DATA for FDR control. The panels from top to bottom show the FDR under weak, moderate and strong annotations, respectively. For each panel, each plot from left to right represents study sample size of 2000, 5000, and 10000, respectively. Within each plot, boxes from left to right represent the proportion of shared risk genes being 0.01, 0.03, 0.05, 0.07 and 0.09, respectively. Each scenario is replicated for 50 times in our simulations.

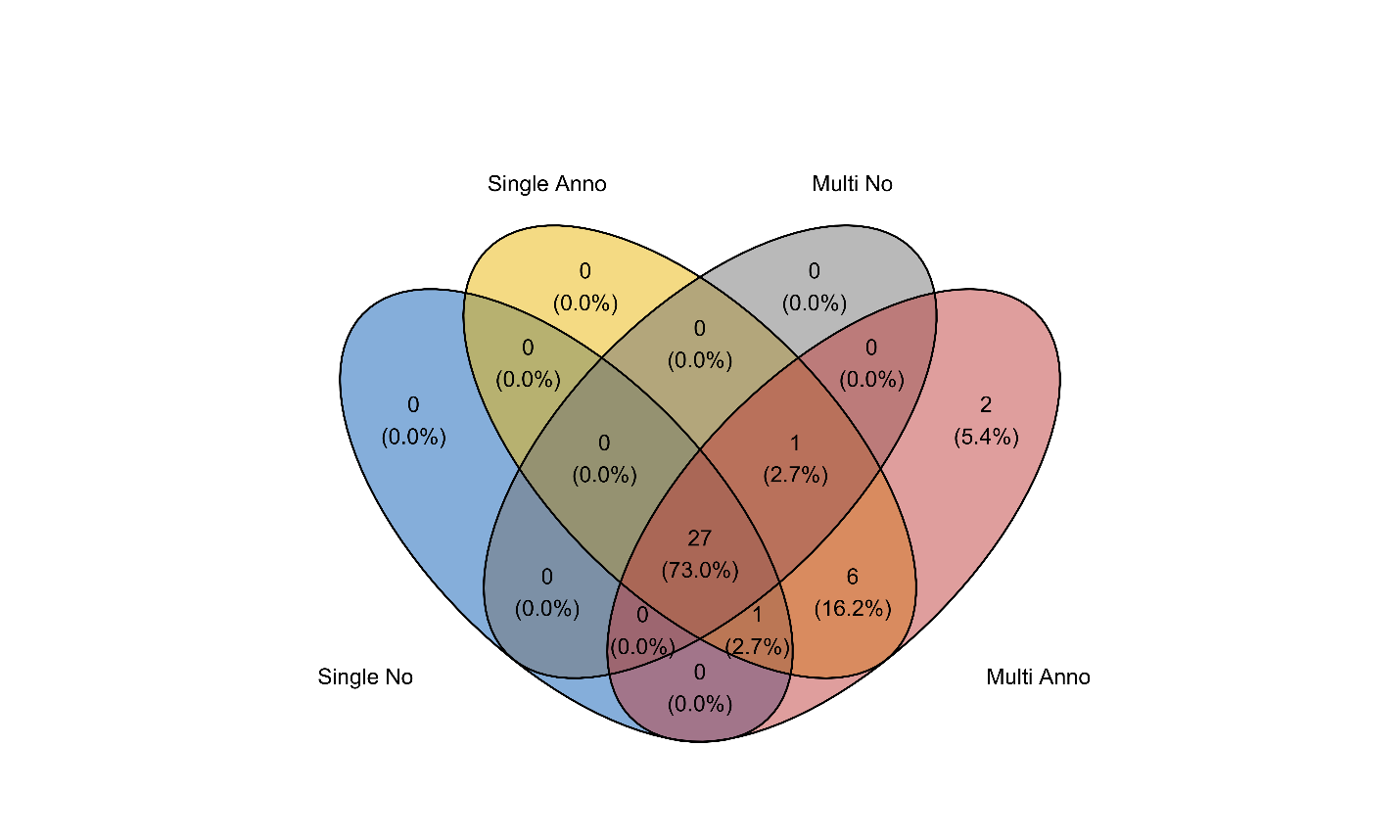

**Fig H**. **Venn diagram of identified genes in different models for autism.** Compared to the single-trait model without annotation, the single-trait model with annotations identified 7 additional genes. Compared to the multi-trait model without annotation, the multi-trait model with annotations identified 9 additional genes. The multi-trait models identified 2 additional genes (*UNC5B* and *BRF1*) compared to the single-trait models.

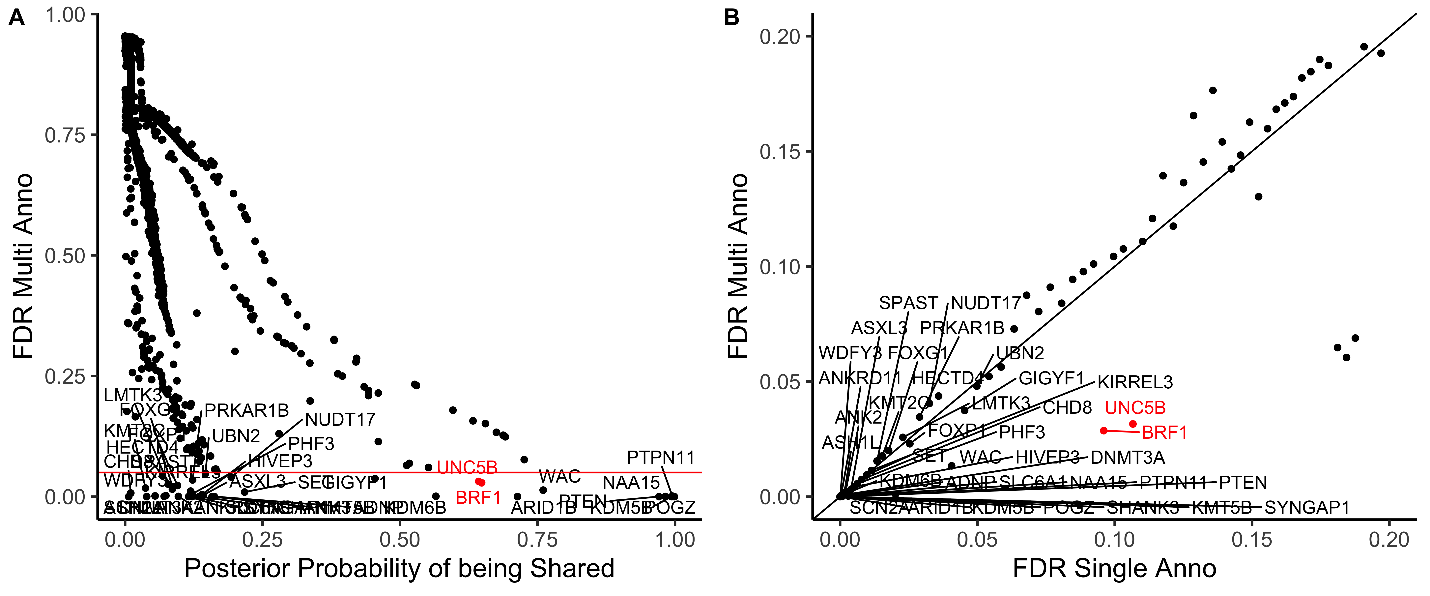

**Fig I. Multi-trait analyses prioritized additional genes with high posterior probability of being shared risk genes for autism.** Gene names of the 37 genes identified by the multi-trait model with annotations are shown on the plot and the additional 2 genes that can be identified by the multi-trait models are marked in red. (A) shows that the 2 additional genes identified by the multi-trait models had high posterior probability of being shared. The x-axis represents the posterior probability of being shared calculated from the multi-trait model with annotations. The y-axis represents the FDR of genes calculated from the multi-trait model with annotations. (B) shows that the top genes in the multi-trait model with annotations also had low FDR (<0.2) in the single-trait model with annotations. The x-axis represents the FDR of genes calculated from the single-trait model with annotations. The y-axis represents the FDR of genes calculated from the multi-trait model with annotations.

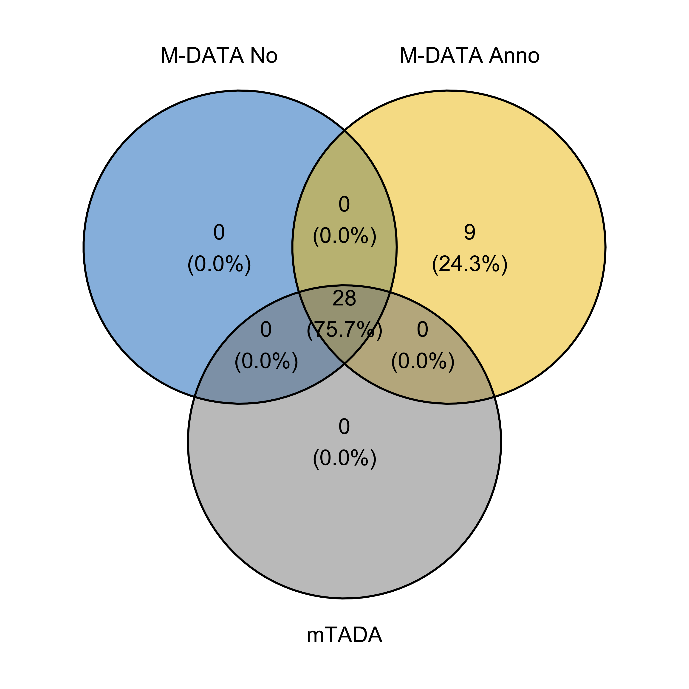

**Fig J. Venn diagram of genes identified by M-DATA and mTADA for autism**. M-DATA multi-trait model with annotations identified 9 additional genes (*BRF1, FOXG1, GIGYF1, HECTD4, LMTK3, NUDT17, SET, UBN2,* and *UNC5B*).
